# Autism-associated MDGA1 missense mutations impair distinct facets of central nervous system development

**DOI:** 10.1101/2025.06.07.25328825

**Authors:** Seungjoon Kim, Hyeonho Kim, Javier Pelayo, Sara Alvarez, Gyubin Jang, Jinhu Kim, Victoria Hoelscher, Beatriz Calleja-Pérez, Hyunsu Jung, Jihae Lee, Seoyeon Kim, Mar Jimenez de la Peña, Ah-reum Han, Dong Sun Lee, Sangho Ji, Wookyung Yu, Ho Min Kim, Joon-Yong An, Won Chan Oh, Seok-Kyu Kwon, Ji Won Um, Alberto Fernández-Jaén, Jaewon Ko

## Abstract

MDGA1 reportedly suppresses GABAergic synaptic inhibition, but it is unclear whether and how MDGA1 dysfunction causes neurodevelopmental disorders. Here, we describe two patients with autism spectrum disorders (ASDs) each carrying pairs of novel missense mutations in *MDGA1*, Val116Met/Ala688Val and Tyr635Cys/Glu756Gln. The Tyr635Cys/Glu756Gln substitution (but not the Val116Met/Ala688V substitution) disrupts the triangular extracellular structure of MDGA1 and renders it unable to impact GABAergic synapses in both cultured hippocampal neurons and hippocampal CA1 pyramidal neurons. Conversely, murine *in utero* overexpression of MDGA1 Val116Met/Ala688Val alters normal cortical neuron migration and impairs ultrasonic vocalizations. Extensive behavioral analyses using forebrain-specific *Mdga1* conditional knockout adult mice revealed a subset of behavioral deficits reminiscent of ASD animal models. Our results collectively demonstrate that different pairs of MDGA1 missense variants associated with ASDs impair distinct facets of central nervous system development via loss-of-function mechanisms.

## Introduction

Mounting evidence from human genetic studies implicates synaptic proteins in the pathogenesis of neuropsychiatric disorders, such as autism spectrum disorders (ASDs), intellectual disability, and schizophrenia (1–3). Efforts have been made to model these so-called synaptopathy-related diseases in various animal species (4, 5). However, it remains difficult to produce appropriate animal models of neuropsychiatric disorders, hindering research efforts to understand the precise mechanisms underlying their pathogeneses and related behaviors (6, 7). A series of converging mechanisms has been associated with ASD-relevant dysfunctions at glutamatergic synapses (1, 8–10), but the detailed molecular and cellular mechanisms underlying GABAergic synaptic dysfunctions remain largely enigmatic.

<u>M</u>eprin, A-5 protein, and receptor protein tyrosine phosphatase mu <u>d</u>omain-containing <u>g</u>lycosylphosphatidylinositol <u>a</u>nchor protein 1 (MDGA1) is a cell surface glycoprotein that attaches to the cell membrane via a glycosylphosphatidylinositol motif (11). MDGA1 functions throughout central nervous system (CNS) development (12, 13) and plays a particular role in regulating cortical neuron migration in the superficial layer (14–17). MDGA1 is also expressed in the adult CNS (18), suggesting that it may contribute to later phases of CNS development or maintenance. Indeed, MDGA1 negatively regulates GABAergic synapse density by binding to synapse organizers, such as neuroligin-2 (Nlgn2) and amyloid precursor protein (APP) (18–20). Intriguingly, the targeting of MDGA1 to APP appears to help tune specific GABAergic neural circuit properties in the hippocampus (20). It remains possible that the Nlgn2-binding activity of MDGA1 contributes to hippocampal synapse organization. Moreover, it is plausible that MDGA1 might deploy additional synaptic pathways in modulating GABAergic synapse development. MDGA1 has been associated with schizophrenia and bipolar disorder (21, 22). Mdga1 knockout (KO) mice exhibited impaired prepulse inhibition of the startle response with altered dopamine and serotonin metabolism (23).

Here, we report two novel missense mutation pairs in the MDGA1 genes of two patients with autistic features, and their functional characterizations in vitro and in vivo. Extensive gain- and loss-of-function analyses in cultured neurons and hippocampal CA1 neurons in mice revealed that expression of the two human MDGA1 variants nullified the ability of wild-type (WT) MDGA1 to regulate neuronal migration and synaptic function. Together with the findings presented in the accompanying paper, our results provide important insights into an MDGA1-associated synaptopathy mechanism underlying ASDs.

## Results

### Clinical manifestations and genetic analyses of two patients with MDGA1 variants

Two patients with autistic features were referred to our clinic. The first patient (P1) was a young girl diagnosed with moderate psychomotor retardation with autistic features by prior clinical and neuropsychological evaluations (Supplemental Table 1). On examination, she showed some mild dysmorphic features, including frontal bossing, prominent eyes, and a thin tented upper lip. The second patient (P2) was a young boy diagnosed with low intellectual functioning and autistic features by prior clinical and cognitive assessments. On examination, he displayed a prominent forehead, blepharophimosis, and thin vermilion of the upper lip. P1 and P2 both lacked relevant family history and had exhibited significant delays in their initial neurodevelopmental milestones. Both cases showed severe delays in verbal and non-verbal communication, as confirmed in their respective neuropsychological evaluations. Routine laboratory screening, neurophysiological tests (electroencephalograms and auditory evoked potentials), and conventional genetic studies (karyotype and aCGH arrays) revealed no significant abnormality.

Whole-exome trio analysis revealed compound heterozygous MDGA1 variants in both cases (Supplemental Table 1). In P1, a paternally inherited missense variant (c.2266G>C; p. Glu756Gln) and a maternally inherited missense variant (c.1904A>G; p. Tyr635Cys) of MDGA1 were identified (Fig. 1A–1D). Both were classified as Variants of Uncertain Significance (VUS) and were found in the Genome Aggregation Database (gnomAD) at extremely low maximal frequencies of 0.07% and 0.001%, respectively, with no healthy homozygote reported. MDGA1 gene had a gnomAD missense Z-Score of 2.57 and the CADD (Combined Annotation-Dependent Depletion) scores of the two mutations were 32 and 27.2, respectively, suggesting that the mutations may have deleterious effects. In P2, a paternally inherited missense variant (c.346G>A; p. Val116Met) and a maternally inherited missense variant (c.2063C>T; p. Ala688Val) of MDGA1 were observed (Fig. 1A–1D). These variants were classified as a VUS and Likely Benign, respectively, and were found in the gnomAD database at extremely low frequencies of < 0.001% and 0.07%, respectively, with no healthy homozygote reported. The CADD scores of these variants were 23.6 and 21.3, respectively, suggesting that they could have deleterious effects. The mutations were observed in the compound heterozygous state in two of P2’s three brothers: One showed mild psychomotor retardation and the other was a very young infant at the time of P2’s evaluation.

**Figure 1.**
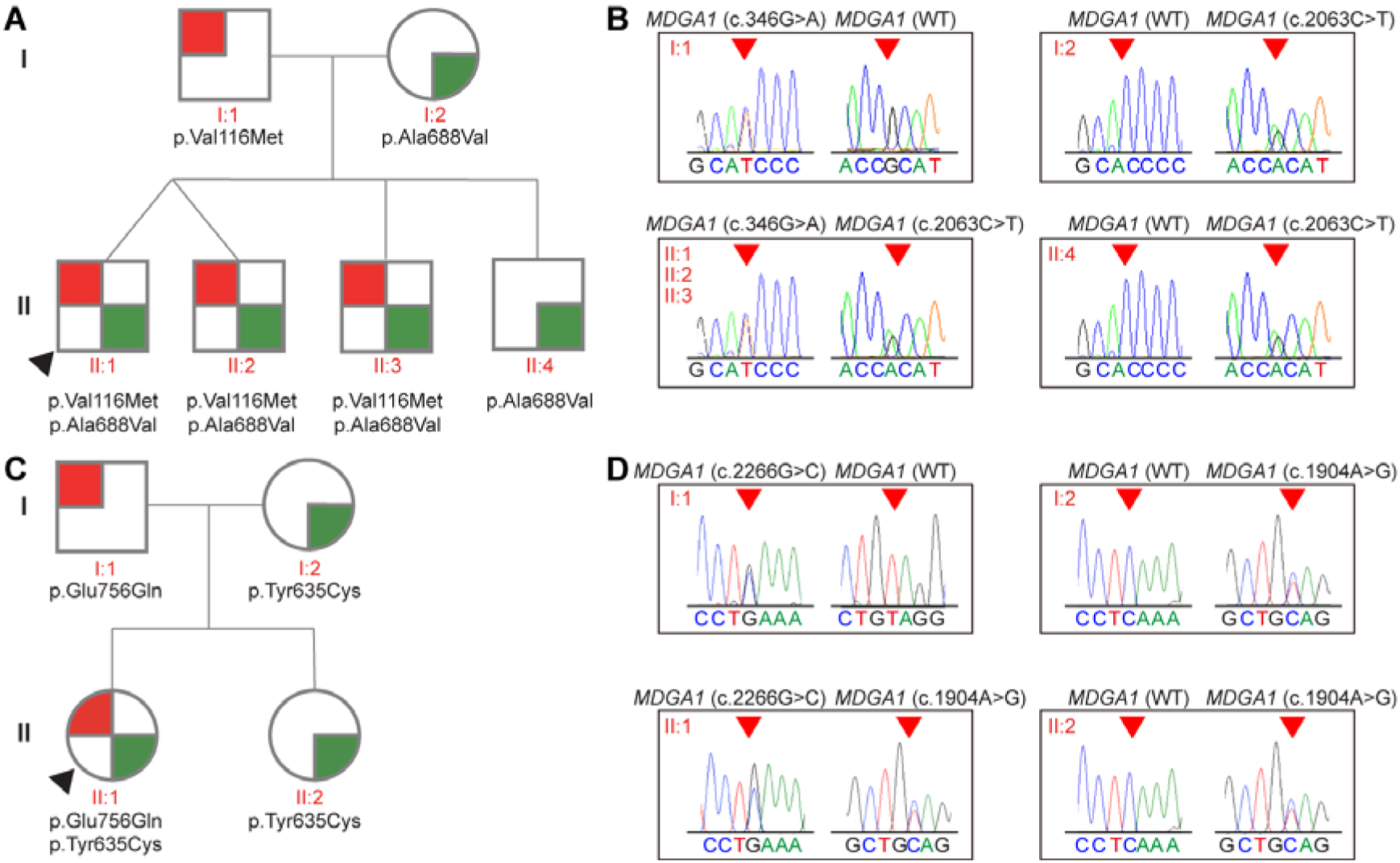
Genetic analysis of human MDGA1 missense mutations found in individuals with autism spectrum disorders. (**A**) Pedigree of a family with MDGA1 (c.346G>A and c.2063C>T) mutations. Squares represent males, circles represent females, and arrows indicate affected individuals. Individuals with the c.346G>A (p.Val116Met) mutation are denoted by red symbols, while those with the c.2063C>T (p.Ala688Val) mutation are denoted by green symbols. (**B**) Sequencing chromatograms showing the MDGA1 (c.346G>A and c.2063C>T) mutations. The chromatograms display the nucleotide the sequences surrounding the mutation sites. (**C**) Pedigree of a family with MDGA1 (c.1904A>G and c.2266G>C) mutations. Symbols and color codes are as described in (**A**). Individuals with the c.1904A>G (p.Glu756Gln) mutation are denoted by red symbols, and those with the c.2266G>C (p.Tyr635Cys) mutation are denoted by green symbols. (**D**) Sequencing chromatograms showing the MDGA1 (c.1904A>G and c.2266G>C) mutations. The chromatograms display nucleotide sequences surrounding the mutation sites.

Sanger sequencing and segregation analysis confirmed that P1 and P2 had inherited the autosomal recessive disorder in a compound heterozygous state, as their mother and fathers were each heterozygous for one the identified variants and were asymptomatic. The WES trio analysis failed to identify any other missense variant with a CADD score over 15 or a loss-of-functional variant with a clear phenotypic association or compatible segregation pattern. Brain 3T MRI analyses of P1 and P2 did not reveal any significant structural malformation (Supplemental Fig. 1). However, in P1, diffusion tensor imaging with 3D-tractography reconstruction showed a marked asymmetry of the superior longitudinal fasciculus (Supplemental Fig. 1).

### Analyses of cellular and temporal expression of MDGA1 in human brain development

To investigate MDGA1 expression throughout brain development, we utilized a single-cell atlas of the human post-mortem brain (24). The atlas collated eight published single-nucleus and single-cell transcriptomic datasets, and unbiasedly integrated data of diverse developmental stages (from gestational week 7 to 90 years) and various cell types (Fig. 2A–2C). Among the 41 clusters of the atlas (Fig. 2C), MDGA1 showed its highest expression levels in Cluster 12 (C12), which comprises excitatory neurons of early cortical development, mainly fetal 2^nd^ trimester cortical excitatory neurons (Fig. 2D). MDGA1 appears to be highly expressed in immature cortical excitatory neurons, particularly from the mid-fetal period to infancy (gestational week 15 to postnatal week 70) (Fig. 2E). We confirmed the excitatory neuron-specific expression of MDGA1 using a previously published single-nucleus atlas (Supplemental Fig. 2A and 2B) (25), which demonstrated a consistently high level of MDGA1 in excitatory neurons (Supplemental Fig. 2C–2F). Specifically, MDGA1 exhibited high-level expression in immature intratelencephalic excitatory neurons and layer II/III intratelencephalic excitatory neurons (Supplemental Fig. 2G). Thus, MDGA1 expression exhibits cellular and temporal specificity in excitatory neurons during the early development of the human cortex.

**Figure 2.**
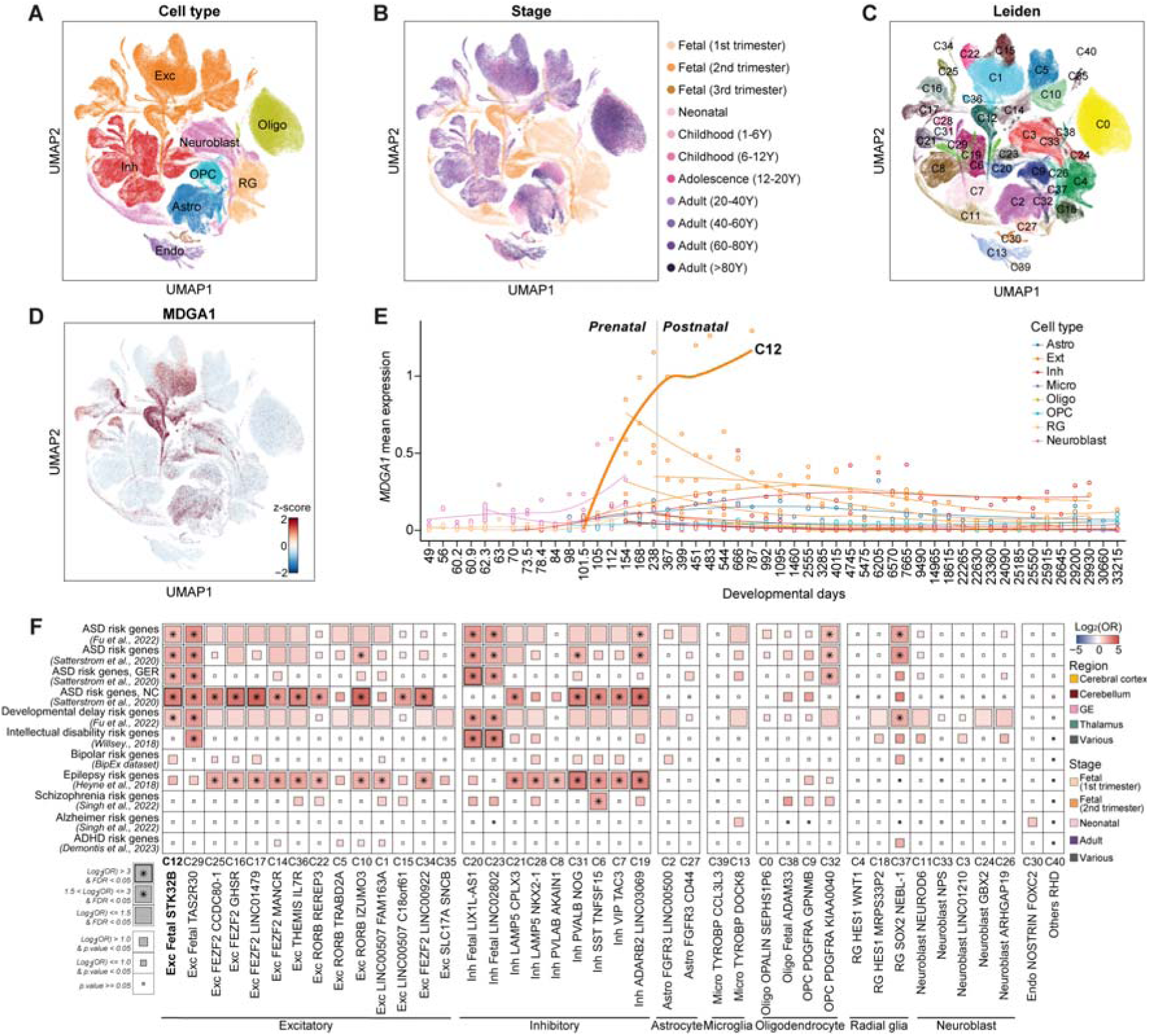
MDGA1 expression in the human brain single-cell atlas. (**A–C**) Uniform Manifold Approximation and Projection (UMAP) of the human brain single-cell atlas, colored according to primary cell types (**A**), developmental stages (**B**), and Leiden cluster (**C**). (**D**) Expression of MDGA1. Colors represent the z-score for MDGA1 expression. (**E**) Expression of MDGA1 over developmental days. The sample-wise mean of log-normalized MDGA1 expression was computed using a pseudobulk method. Clusters with at least 4,600 cells (C0-C22) were used. (**F**) Gene set enrichment test for neurological disorder genes. Fisher exact test was used to compute statistics. Significant tests were defined as those having an adjusted *p*-value < 0.05 and an odds ratio (log2) > 1.5

Given that risk genes for ASDs and neurodevelopmental disorders are reportedly enriched during early cortical neurodevelopment (26, 27), we performed gene set enrichment tests for neurological genes among the cluster-specific genes. We found that the C12-specific genes (n = 1,632) were significantly enriched for ASD and developmental delay risk genes (False Discovery Rate < 0.05; Fisher’s exact test). Additionally, C12 was robustly associated with bipolar risk genes and epilepsy risk genes (Log_2_(OR) > 1, p < 0.05). These observations collectively suggest that MDGA1 may play a role across a spectrum of neuropsychiatric conditions with a shared genetic underpinning (Fig. 2F; Supplemental Table 2). Notably, many of the C12-specific ASD-risk genes (11/24) were previously reported by Satterstrom et al. (26) as being neuronal communication genes involved in ASD risk, potentially suggesting that MDGA1 expressed in projection neurons is relevant to early synaptogenesis. We also found that the expression of MDGA1 was increased during the fetal to postnatal transition period, which is consistent with its proposed role as a neuronal communication gene in ASD risk (26).

### Normal expression level, surface transport, ligand-binding, and folding of MDGA1 in the MDGA1 mutations of P1 and P2

To investigate how the MDGA1 mutations identified in P1 and P2 impacted the protein structure of human MDGA1 (hMDGA1), we predicted the structure of hMDGA1 using AlphaFold2 (28, 29). We found that, within hMDGA1, V116M resides within the Ig1 domain, Y635C and A688V are found in its fibronectin type III (FNIII) domain, and E756Q localizes to the MAM domain (Fig. 3A). Since the structure of the MAM domain was not included in the published X-ray crystallographic results for MDGA1 (30), we were unable to predict any structural alteration related to the E756Q mutation. Regarding the V116M mutation, it is known that hMDGA1 and neuroligin-2 (Nlgn2) form a hetero-tetrameric complex that exhibits a 2:2 stoichiometry and is strongly connected via a long-range of electrostatic interactions and hydrogen bonds (31). At amino acid (aa) 116 of MDGA1, which is located at the binding interface, the replacement of a small nonpolar valine with a long methionine (V116M), is likely to disrupt interactions between the Ig1 domain of MDGA1 and Nlgn2 via steric hindrance with neighboring polar residues (R243 and E247) without significantly altering the 3D structure of MDGA1 (Fig. 3A). Regarding the Y635C mutation, the previously solved crystal structure of chicken MDGA1 (Ig_1-6_-FNIII) showed that it takes on a triangular shape in which the sharply angled Ig_2_-Ig_3_, Ig_3_-Ig_5_ and Ig_6_-FNIII linkers are stabilized by multiple inter-domain contacts (30). hMDGA1 Y635, corresponding to chicken MDGA1 Y629, is located at the interface between Ig6 and FNIII (Supplemental Fig. 3). Therefore, we hypothesized that the hMDGA1 Y635C mutation could structurally weaken these inter-domain interactions, as supported by our free energy calculations (Supplemental Table 3), and thereby disrupt the triangular structure of hMDGA1. Our negative-stain electron microscopy confirmed the structural prediction (30) by showing that hMDGA1 WT exists as a closed triangular shape (blue box). In contrast, most of the hMDGA1 Y635C/E756Q mutant particles exhibited linear or irregular structures (red box) (Supplemental Fig. 4A). Finally, it is likely that the A688V mutation in the FNIII domain of hMDGA1 might mildly affect the protein stability of the FNIII domain.

**Figure 3.**
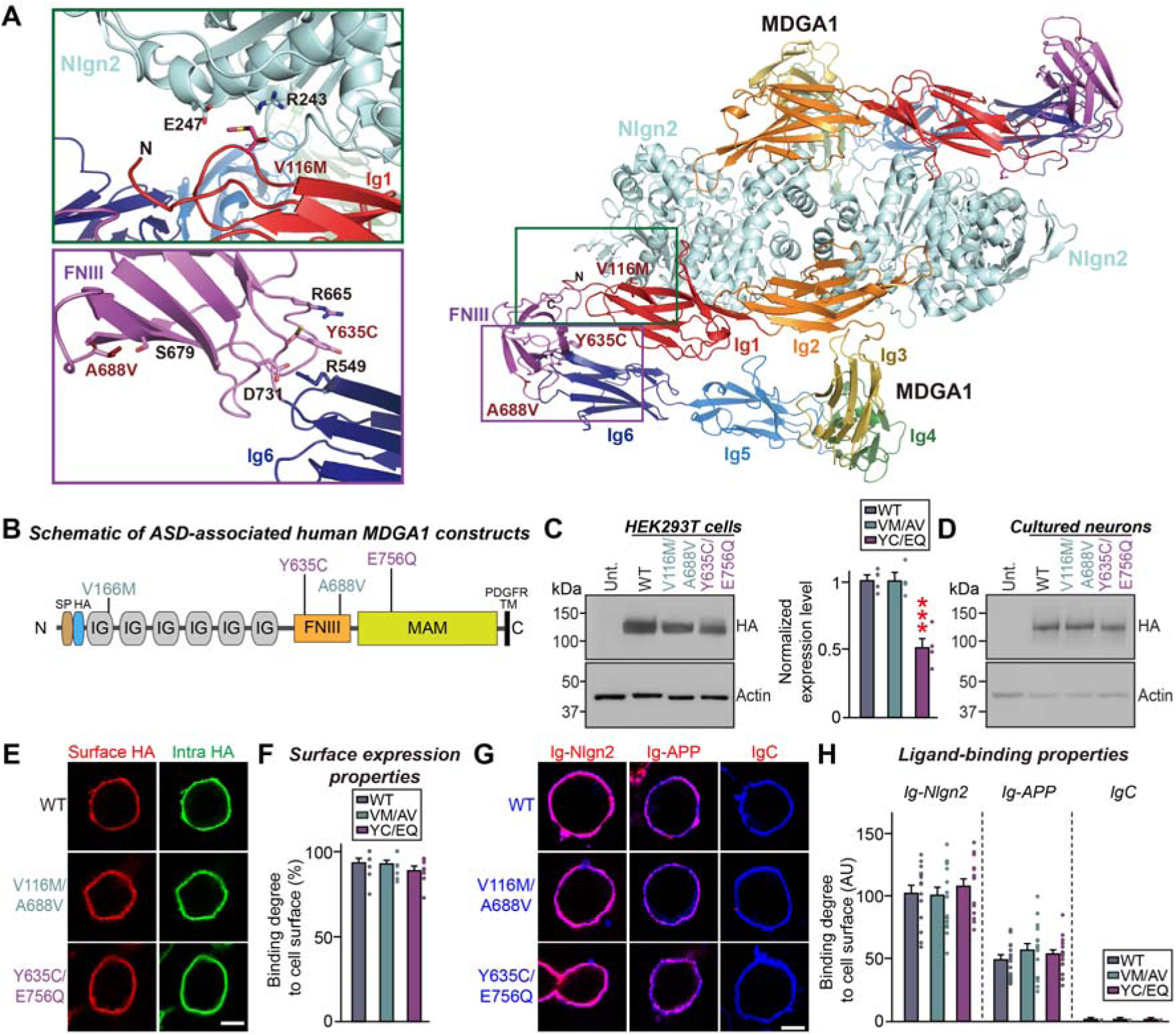
Biochemical and cellular properties of the autism spectrum disorder-linked human MDGA1 missense mutations. (**A**) Structural model of the hetero-tetrameric human MDGA1/neuroligin complex (PDB: 5OJ6; PDB: 5XEQ)(30, 31) and patient mutations in human MDGA1. For clarity, only the crystal structure of Nlgn2 and the aligned structural model of human MDGA1 are presented. Domains of human MDGA1 (six Ig domains and one FnIII domain) are colored with a rainbow gradient from Ig1 (red) to FnIII (violet). Dimeric Nlgn2 is colored with light cyan. Close-up views of mutated and neighboring residues in human MDGA1 (**left**). Residues corresponding to patients’ mutations described in the current study are presented as sticks and labeled. (**B**) Diagram illustrating the ASD-associated MDGA1 variants used in the current study. (**C and D**) Immunoblot analyses probing the expression levels of MDGA1 WT and the indicated variants in HEK293T cells (**C, left**) or cultured hippocampal neurons (**D**). Quantification of the expression levels of the indicated MDGA1 variants (**C**, **right**) was presented. Data are means ± SEMs (‘n’ denotes number of independent experiments; n = 4 for all experimental groups; \*\*\**p* < 0.001; Tukey’s multiple comparison test). (**E and F**) Analyses of the surface transport activity of HA-tagged MDGA1 WT and its variants in HEK293T cells. Representative images (**E**) of transfected HEK293T cells, illustrating the expression and surface transport activity of MDGA1 variants. Transfected cells were fixed (not permeabilized) and incubated with anti-HA antibody to detect the extracellular region of MDGA1 (red). Intracellular levels of MDGA1 constructs were visualized by incubating with an anti-HA antibody after permeabilization (green). Scale bar, 10 μm (applies to all images). Quantification (**F**) of surface transport activity of MDGA1 constructs. Data are means ± SEMs (nonparametric Kruskal-Wallis test with Dunn’s *post hoc* test; n = 10 images/group). Scale bar, 10 μm (applies to all images). (**G** and **H**) Cell surface-binding assays. Representative images (**G**) showing HEK293T cells expressing N-terminally HA-tagged MDGA1 WT and its variants incubated with purified Ig-fused neuroligin-2 (Ig-Nlgn2), Ig-fused APP (Ig-APP) or IgC alone (control), analyzed by immunofluorescence imaging for Ig-fusion proteins (red) and HA (blue). Scale bars, 10 μm (applies to all images). Quantification of cell surface binding (**H**). Data are means ± SEM (n = 12–18 cells/group). Scale bar, 10 μm (applies to all images).

To next examine whether the ASD-linked MDGA1 mutants exhibit altered secondary structures, we measured circular dichroism (CD) spectra. We observed a strong negative peak near 210 nm for MDGA1 wild type (WT), suggesting that its main secondary structure is composed of β- sheets. The CD spectra of MDGA1 V116M/A688V and MDGA1 Y635C/E756Q were similar to that of MDGA1 WT (Supplemental Fig. 4B). Thus, the ASD-linked MDGA1 mutations did not appear to alter the secondary structures of MDGA1.

To further probe the impact of the identified a.a. substitutions on the structure and/or synaptic function of MDGA1, we generated mammalian expression vectors encoding V116M/A688V and Y635C/E756Q variants of MDGA1, both of which were identified as potentially damaging by various in silico prediction tools (Supplemental Table 1). Regarding conservation, we note that Y635 and E756 of hMDGA1 are conserved at equivalent positions in hMDGA2, and V116, Y635, and E756 (but not A688) of hMDGA1 are largely conserved across various vertebrate species and thus may be functionally significant (Supplemental Fig. 2).

To compare the expression levels and intracellular transport properties of the MDGA1 variants versus MDGA1 WT, we expressed HA-tagged variants and WT MDGA1 in human embryonic kidney 293T (HEK293T) cells. Immunoblot analyses of lysates from HEK293T cells or cultured hippocampal neurons showed that total protein expression levels of MDGA1 V116M/A688V or MDGA1 Y635C/E756Q were comparable to those of MDGA1 WT (Fig. 3B–3D). Analysis of the surface and intracellular protein levels of MDGA1 WT and the mutants in HEK293T cells revealed that MDGA1 V116M/A688V or MDGA1 Y635C/E756Q exhibited surface levels similar to those of MDGA1 WT (Fig. 3E and 3F). To determine whether the identified mutations affected the interactions of MDGA1 with known extracellular ligands (Nlgn2 or APP), we assayed the cell-surface binding of recombinant Ig-fusion proteins of MDGA1 (IgC-MDGA1) or IgC alone (negative control) with HEK293T cells expressing HA-tagged Nlgn2 or APP. Our results revealed that IgC-MDGA1 proteins robustly bound to HEK293T cells expressing HA-Nlgn2 or HA-APP, whereas IgC did not (Fig. 3G and 3H). We attempted to analyze whether the MDGA1 mutations could alter the synaptic localization of MDGA1. However, in line with a prior observation (32, 33), overexpression of MDGA1 WT and its variants in cultured mature hippocampal neurons yielded diffuse distribution along the dendrites.

### Overexpressed MDGA1 Y635C/E756Q fails to decrease GABAergic synapses in cultured hippocampal neurons

Given previous reports that MDGA1 overexpression specifically reduces the number of GABAergic synapses in cultured hippocampal neurons and hippocampal CA1 pyramidal neurons (18–20), we explored whether our ASD-associated MDGA1 mutations could impair the ability of MDGA1 to alter GABAergic synapses. To this end, we transfected the indicated MDGA1 variants into cultured hippocampal neurons at days in vitro 7 (DIV7) and measured the density of puncta positive for both gephyrin (a marker for postsynaptic GABAergic synaptic specialization) and vesicular GABA transporter (VGAT; a marker for presynaptic GABAergic nerve terminals) at DIV14 (Fig. 4A and 4B). In line with previous reports (18, 19), overexpression of MDGA1 WT significantly decreased the density and area of gephyrin^+^VGAT^+^ puncta. Interestingly, overexpressed MDGA1 V116M/A688V had similar effects, whereas overexpressed MDGA1 Y635C/E756Q failed to alter the GABAergic synapse puncta density or area (Fig. 4A and 4B). However, both ASD-associated MDGA1 variants effectively suppressed Nlgn2-induced presynaptic assembly in heterologous synapse formation assays, in a manner similar to MDGA1 WT (Supplemental Fig. 5). Further experiments showed that the E756Q substitution, but not the Y635C substitution, was responsible for abrogating the activity of MDGA1 WT in increasing the GABAergic synapse number in cultured neurons (Supplemental Fig. 6). Since MDGA2 overexpression also increases excitatory synapses (33), we questioned whether overexpression of MDGA2 Y631C/E751Q also fails to suppress excitatory synapse density. However, overexpressed MDGA2 Y631C/E751Q and MDGA2 WT displayed similar abilities to suppress excitatory synapses (Supplemental Fig. 7A and 7B).

**Figure 4.**
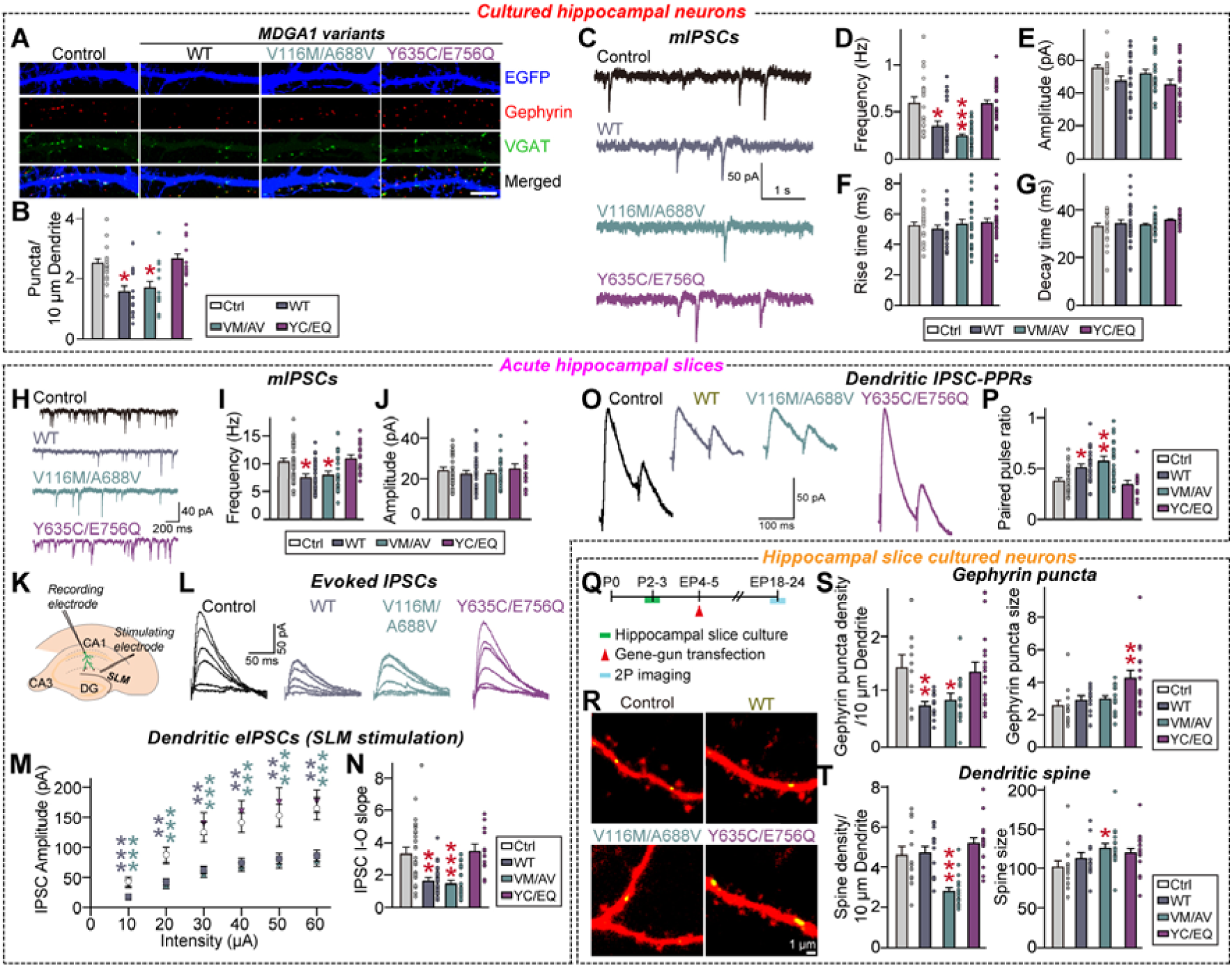
Analysis of GABAergic synaptic properties in hippocampal CA1 pyramidal neurons expressing ASD-associated MDGA1 variants. (**A** and **B**) Representative images (**A**) and summary graphs (**B**) showing the density of synaptic puncta in cultured hippocampal neurons transfected at DIV7 with the indicated full-length MDGA1 expression construct and immunostained at DIV14 with antibodies to VGAT, gephyrin and EGFP. Data are presented as means ± SEMs (n = 14–15 images/group; \**p* < 0.05; ANOVA with non-parametric Kruskal-Wallis test). Scale bar, 10 μm (applies to all images). (**C–G**) Representative mIPSCs traces (**C**) and quantification of frequency (**D**), amplitude (**E**), rise time (**F**), and decay time (**G**) of mIPSCs from cultured neurons transfected with MDGA1 WT or its variants. Data are presented as means ± SEMs (control, n = 19; WT, n = 22; V116M/A688V, n = 25; Y635C/E756Q, n = 24; \**p* < 0.05, \*\*\**p* < 0.001; ANOVA with a non-parametric Kruskal-Wallis test). (**H–J**) Representative traces (**H**) of electrophysiological recordings in CA1 pyramidal neurons expressing MDGA1 WT and its variants. Quantification of mIPSC frequency (**I**) and amplitude (**J**) recorded in the hippocampal CA1 pyramidal neurons. Data are presented as means ± SEMs (n = 16– 37/group; \**p* < 0.05; ANOVA with non-parametric Kruskal-Wallis test). (**K–N**) Experimental setup for recording eIPSCs in hippocampal CA1 pyramidal neurons by placing stimulating electrodes in the *stratum lacunosum-moleculare* (SLM) layer. Representative traces (**L**) of eIPSCs recorded from dendritic recordings in CA1 pyramidal neurons. Average eIPSC I-O curve (**M**), and average eIPSC I-O slope (**N**) from hippocampal CA1 pyramidal neurons expressing the indicated MDGA1 variants. Data are presented as means ± SEMs (n =12–27/group; \*\**p* < 0.01, \*\*\**p* < 0.001, ANOVA with non-parametric Kruskal-Wallis test). (**O** and **P**) Representative somatic eIPSC paired-pulse ratio (PPR) traces (**P**) and average PPR (**Q**) from hippocampal CA1 pyramidal neurons expressing the indicated MDGA1 variants. Data are presented as means ± SEMs (n = 12–27/group; \**p* < 0.05, \*\**p* < 0.01; ANOVA with non-parametric Kruskal-Wallis test). (**Q**) Schematic showing biolistic transfection of organotypic hippocampal CA1 slices with MDGA1 mutants at EP4–5 and imaging at EP18–24. (**R**) Two-photon images of dendritic segments from CA1 pyramidal neurons co-transfected with tdTomato, gephyrin intrabody-GFP, and MDGA1 control, MDGA1 WT, MDGA1 V116M/A688V, or MDGA1 Y635C/E756Q vector. (**S** and **T**) Quantitative analysis of gephyrin puncta density and size (**S**) and dendritic spine density and size (**T**) across conditions.

To test whether the ASD-associated MDGA1 mutations could impact the MDGA1-mediated negative regulation of GABAergic synaptic transmission, we measured miniature inhibitory postsynaptic currents (mIPSCs) using whole-cell voltage clamp recordings. Overexpression of MDGA1 WT significantly decreased the mIPSC frequency without altering the mIPSC amplitude, rise time, or decay time, compared to those from neurons expressing control plasmid (Fig. 4C–4G).

Overexpressed MDGA1 V116M/A688V had a similar effect, whereas overexpressed MDGA1 Y635C/E756Q did not alter GABAergic synaptic transmission (Fig. 4C–4G). However, overexpression of MDGA2 Y631C/E751Q significantly decreased the frequency of miniature excitatory postsynaptic currents (mEPSCs), in keeping with the two-photon imaging results (Supplemental Fig. 7C–7G).

Taken together, these data suggest that the MDGA1 Y635C/E756Q substitutions manifest as an MDGA1 loss-of-function mutation in the regulation of GABAergic synaptic properties.

### Overexpressed MDGA1 Y635C/E756Q fails to negatively regulate GABAergic synaptic properties in hippocampal CA1 pyramidal neurons

We recently showed that overexpression of MDGA1 in the adult mouse hippocampal CA1 suppresses GABAergic synaptic transmission and stabilization (20). To determine whether our ASD-associated MDGA1 mutations could affect the ability of MDGA1 to negatively regulate the GABAergic synapse properties, we injected adeno-associated viruses (AAVs) expressing MDGA1 WT or the indicated MDGA1 variants into the hippocampal CA1 of adult mice and performed whole-cell patch clamp recordings to measure the GABAergic synaptic transmission and synaptic strength.

Consistent with our previous report (20), overexpression of MDGA1 WT significantly decreased the frequency (but not amplitude) of mIPSCs in the hippocampal CA1 pyramidal neurons (Fig. 4H–4J). Overexpression of MDGA1 V116M/A688V had a similar effect, whereas that of MDGA1 Y635C/E756Q failed to decrease the mIPSC frequency (Fig. 4H–4J). To examine whether MDGA1 has a similar impact on GABAergic synaptic transmission in the medial prefrontal cortex (mPFC), we measured mIPSCs in layer II/III of mPFC pyramidal neurons. Surprisingly, overexpression of MDGA1 WT and its ASD-associated variants did not alter the frequency or amplitude of mIPSCs nor the densities of gephyrin puncta or dendritic spines (Supplemental Fig. 8).

We next examined whether the ASD-associated MDGA1 mutations impacted the ability of MDGA1 to suppress evoked inhibitory postsynaptic currents (eIPSCs) in the distal dendritic compartment of hippocampal CA1 pyramidal neurons (20). We found that overexpression of MDGA1 WT or MDGA1 V116M/A688V significantly decreased the amplitude of distal dendritic eIPSCs and increased the paired pulse ratio (PPR), whereas MDGA1 Y635C/E756Q failed to do so (Fig. 4K–4P). In contrast, overexpression of MDGA1 WT, V116M/A688V, or Y635C/E756Q had no effect on the frequency or amplitude of asynchronous IPSCs (aIPSCs) (Supplemental Fig. 9).

To corroborate our electrophysiological observations, we used two-photon laser-scanning microscopy to examine gephyrin puncta in hippocampal CA1 slice cultures (Fig. 4Q). Our results revealed that overexpression of MDGA1 WT or MDGA1 V116M/A688V significantly decreased the gephyrin puncta density, whereas overexpression of MDGA1 Y635C/E756Q did not; in fact, overexpression of the latter variant tended to increase the gephyrin puncta size (Fig. 4R and 4S).

Unexpectedly, overexpression of MDGA1 V116M/A688V, but not MDGA1 WT or MDGA1 Y635C/E756Q, significantly reduced the density of dendritic spines while concomitantly increasing their size (Fig. 4R and 4T). These results were further reinforced by data obtained from cultured hippocampal neurons, wherein overexpression of MDGA1 V116M/A688V, but not MDGA1 WT or MDGA1 Y635C/E756Q, specifically decreased the excitatory synapse density and mEPSC frequency (Supplemental Fig. 10). Our findings collectively indicate that the Y635C/E756Q substitution renders MDGA1 unable to control GABAergic synapse organization in the hippocampal CA1, whereas the MDGA1 V116M/A688V substitution affects excitatory synaptic structures.

### In vivo overexpression of MDGA1 V116M/A688V, but not MDGA1 Y635C/E756Q, alters cortical neuron migration and ultrasonic vocalizations in mice

In patients with ASD, the marked neuropathological characteristics include defective neuronal migration (34). Intriguingly, MDGA1 knockdown in migrating layer II/III cortical neurons can disrupt their normal migration(14). Moreover, transient glutamatergic synapse formation between subplate neurons and immature migratory neurons instructs neocortical neuronal migration (35), reminiscent of the excitatory synaptic phenotype of MDGA1 V116M/A688V (see Figs. 4 and Supplemental Fig. 10). To investigate whether our ASD-associated MDGA1 variants could alter cortical neuron migration, we performed in utero electroporation experiments. Expression plasmids encoding HA-tagged MDGA1 (WT, V116M/A688V or Y635C/E765Q) or a control plasmid (pCIG2 harboring an internal ribosome entry site-driven enhanced green fluorescent protein [EGFP] sequence) were electroporated into mice at E15.5 to mainly target progenitors of layer II/III cortical pyramidal neurons, but not GABAergic interneurons (Fig. 5A), as previously described (36). At P21, neuronal positions in the electroporated mice were examined by EGFP fluorescence (Fig. 5A).

**Figure 5.**
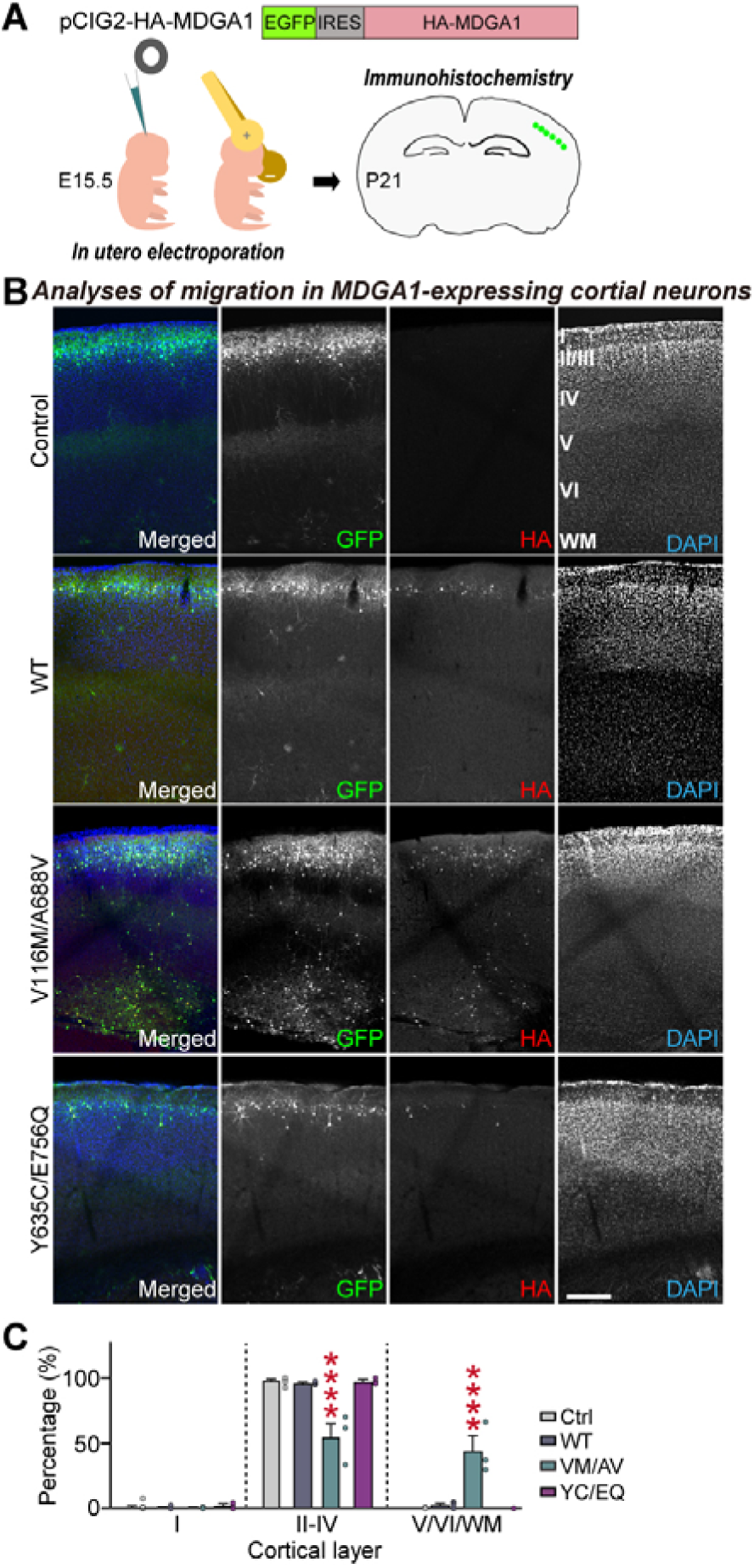
Analysis of cortical neuronal migration in mice expressing ASD-associated MDGA1 variants. (**A**) Schematic of the *in utero* electroporation. The indicated plasmids (control or HA-tagged MDGA1 variants [HA-MDGA1-IRES-EGFP]) were electroporated *in utero* at E15.5 and neuronal migrations was analyzed by immunohistochemistry using confocal microscopy at P21. (**B** and **C**) Analysis of the migration trajectory of cortical neurons expressing the indicated MDGA1 variants. Data are presented as means ± SEMs (‘n’ denotes number of images/mice; control, n = 18/9; WT, n = 10/5; V116M/A688V, n = 6/3; and Y635C/E756Q, n = 5/3; \*\*\*\**p* < 0.0001; Mann Whitney *U* test). Scale bar, 500 nm.

Neurons expressing MDGA1 WT or MDGA1 Y635C/E756Q exhibited patterns of migration across cortical layers similar to those of neurons expressing the control plasmid (> 95% neurons reached layers II/III) (Fig. 5B and 5C). Strikingly, neurons expressing MDGA1 V116M/A688V displayed abnormally altered migration: Only ∼55% of the transfected neurons were seen in layer II/III, and a large portion were detected in layer V/VI/white matter (∼44.5%) (Fig. 5B and 5C). These results suggest that the V116M/A688V substitution might have a dominant-negative inhibitory effect on MDGA1 function, leading to abnormal cortical neuron migration. These findings further suggest that overexpression of different ASD-associated MDGA1 variants induces distinct MDGA1 loss-of-function effects in vivo. Indeed, and in line with a previous report (14), MDGA1-deficient cortical neurons from Mdga1 floxed mice crossed with Emx1-Cre mice (Emx1-Cre::Mdga1^f/f^, also denoted as Mdga1-cKO) exhibited abnormal migration patterns, as assessed via immunohistochemistry using antibodies against layer-specific markers (Supplemental Fig. 11). Since MDGA1 is primarily expressed in excitatory neurons of the cortex and hippocampus of adult mice (18, 37), crossing Mdga1 floxed mice with Nestin-Cre failed to yield enough animals for in-depth analyses (data not shown), and human MDGA1 is profusely expressed in excitatory neurons (see Fig. 2), the forebrain excitatory neuron-specific Mdga1-cKO mice were used for the remaining experiments, as also employed in our prior study (20).

### Mdga1-cKO mice exhibit various autistic-like abnormal behaviors

Previous studies showed that constitutive Mdga1-KO mice exhibit impaired hippocampus-dependent spatial learning and memory and deficiency in the startle response (23, 37), which are typically considered to be schizophrenia endophenotypes (38). In view of the observation that P1 and P2 patients harboring the MDGA1 missense mutations commonly exhibit communication abnormalities, we evaluated maternal isolation-induced ultrasonic vocalizations (USVs), which are distress calls emitted by pups when separated from their mother (representing infant-mother vocal communicative behavior relevant to ASD) (39). In utero electroporation was performed for control, MDGA1 WT, MDGA1 V116M/A688V, or MDGA1 Y635C/E756Q vectors, and maternal isolation-induced USVs of pups were recorded on postnatal days 3 (P3), P6, P9, and P12. Call numbers in all pups increased from P3 to P6, decreased slightly by PND9, and reached zero by P12 (Supplemental Fig. 12A and 12B). Intriguingly, MDGA1 V116M/A688V-expressing mice emitted significantly fewer USVs compared to age-matched pups expressing control, MDGA1 WT, or MDGA1 Y635C/E756Q at all ages (up to P12) (Supplemental Fig. 12A and 12B).

To further establish whether the MDGA1 V116M/A688V-induced impairment of isolation-induced USVs reflects gain- or loss-of-function changes in MDGA1, we examined isolation-induced USVs in Mdga1-cKO pups (Supplemental Fig. 12C and 12D). These pups exhibited USV deficits on P6 and P9 similar to those seen in MDGA1 V116M/A688V-expressing pups. The Mdga1-cKO pups emitted fewer USVs when separated from mothers during PND3–PND12 (Supplemental Fig. 12C). Regarding other autistic-like behavioral phenotypes (see Fig. 6A for a schematic showing the flow of behavioral assays), male adult (P60) Mdga1-cKO mice exhibited levels of anxiety/exploration-related behavior, locomotor activity, working memory, and repetitive/compulsive-like behavior within the expected ranges (Supplemental Fig. 13). However, adult male Mdga1-cKO mice exhibited deficits in social behavior, as evident from their reduced time spent exploring animated mice over an object (sociability) and reduced time spent exploring unfamiliar mice (social novelty recognition) in the three-chamber test (Fig. 6B–6D). These mice also emitted reduced USVs upon encountering a female mouse, as well as a shift in the distribution of syllable types in the USVs (Fig. 6E–6H). Impaired sensorimotor gating was observed in adult male Mdga1-cKO mice, as characterized by decreases in the acoustic startle response and prepulse inhibition (Fig. 6I–6K), as well as decreased novel object-recognition memory (Fig. 6L–6N). The home-cage activities of these mice were unaltered (Fig. 6O–6T).

**Figure 6.**
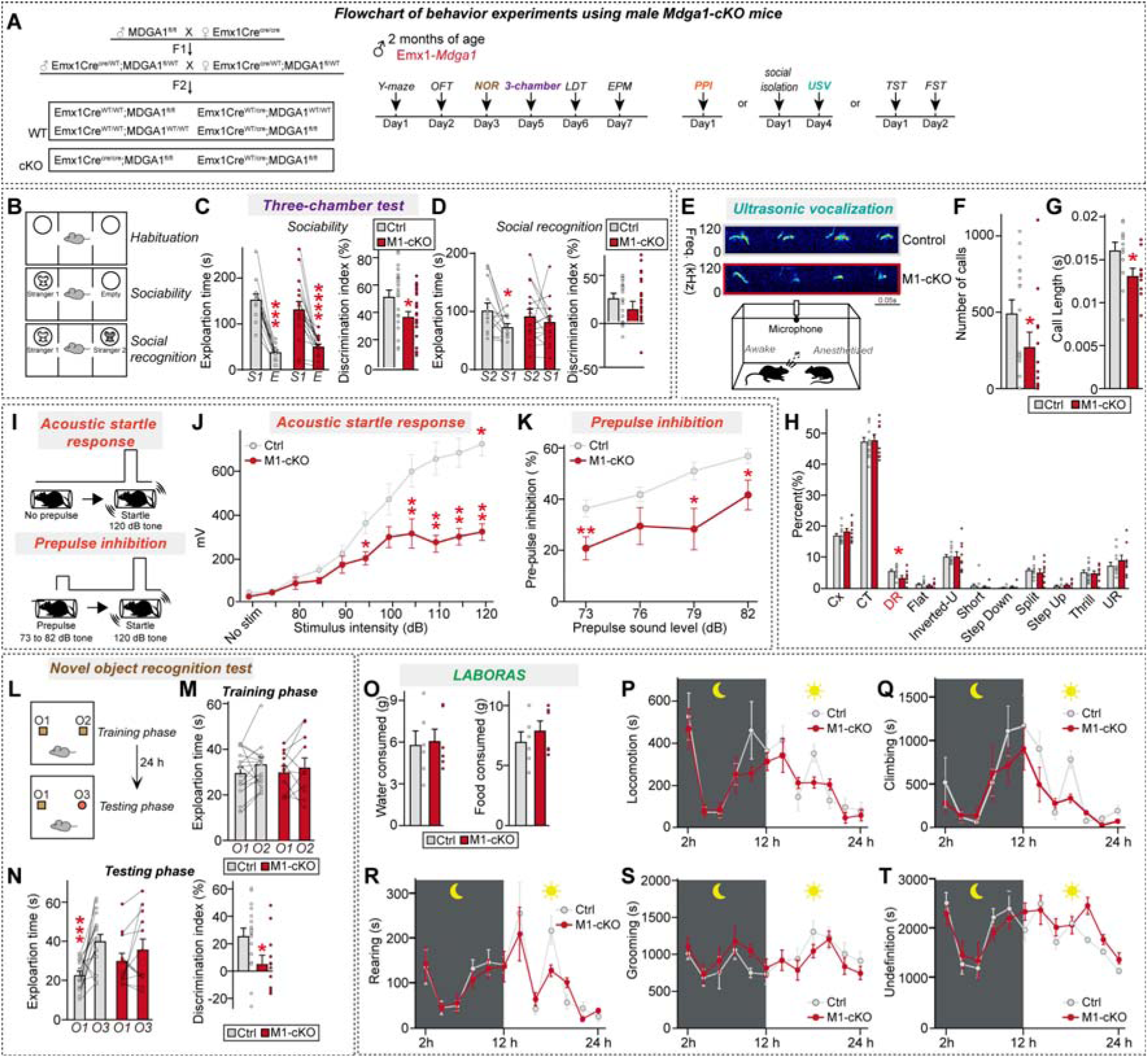
Analysis of mouse behavior in male adult *Mdga1*-cKO mice. (**A**) Breeding scheme followed to generate *Mdga1*-cKO mice. *Emx1*-Cre driver mice were crossed with *Mdga1*-floxed mice. The timeline of behavioral experiments performed on male *Mdga1*-cKO mice, including Y-maze, open field test (OFT), novel object recognition (NOR), three-chamber, light-dark transition (LDT), elevated plus maze (EPM), tail suspension test (TST), forced swim test (FST), social isolation, USV, and prepulse inhibition (PPI) tests. (**B–D**) Schematic (**B**) of the three-chamber social interaction test used to assess sociability and social recognition in mice. Quantification of exploration time during the three-chamber test for sociability (**C**; stranger 1 vs. empty) and social recognition (**D**; stranger 1 vs. stranger 2) in adult male control and *Mdga1*-cKO mice. Data are presented as means ± SEMs (n = 13–16 mice/group; \**p* < 0.05, \*\*\**p* < 0.001, \*\*\*\**p* < 0.0001; Wilcoxon matched-pairs signed rank test). (**E–H**) Representative USV traces and schematic (**E**) of the recording setup for adult mice. Number of USV calls (**F**) and USV call length (**G**) recorded from adult male control and *Mdga1*-cKO mice. Data are presented as means ± SEMs (n = 10–14 mice/group; \**p* < 0.05; Mann–Whitney *U* test). Percent distribution of different USV call types (**H**; complex trill, downward ramp, flat, inverted-U, short, split, step down, step up, trill, upward ramp) in adult male control and *Mdga1*-cKO mice. Data are presented as means ± SEMs (n = 12–15 mice/group; \**p* < 0.05; Mann–Whitney *U* test). (**I–K**) Schematic (**I**) of the PPI test. Mice were exposed to a prepulse sound (73 to 82 dB) followed by a startle pulse (120 dB), and the inhibition of the startle response was measured. Acoustic startle response (**J**) in adult male control and *Mdga1*-cKO mice. Data are presented as means ± SEMs (n = 12– 18 mice/group; \**p* < 0.05, \*\**p* < 0.01, Mann–Whitney *U* test). Prepulse inhibition of the acoustic startle response (**K**) in adult male control and *Mdga1*-cKO mice. Data are presented as means ± SEMs (n = 12–18 mice/group; \**p* < 0.05, \*\**p* < 0.01, Mann–Whitney *U* test). (**L–N**) Schematic (**L**) of the NOR test. Training phase and testing phase timelines are shown, with object placements (O1, O2, O3) indicated. Quantification of exploration time during the training phase (**M**) and testing phase and the discrimination index (**N**) of the NOR test in adult male control and *Mdga1*-cKO mice. Data are presented as means ± SEMs (n = 12–18 mice/group; \**p* < 0.05, \*\*\**p* < 0.001; Wilcoxon matched-pairs signed rank test). (**O–T**) Quantification of various homecage activities in LABORAS cages, where mouse movements were monitored for 24 h. Shown are water and food consumption in grams (**O**), and locomotion (**P**), climbing (**Q**), rearing (**R**), grooming (**S**), and undefined (**T**) behaviors of adult male control and *Mdga1*-cKO mice. Data are plotted every 2 h. The light and dark cycles of the day are represented by sun and moon icons, respectively. Data are presented as means ± SEMs (n = 6 mice/group).

Given that there is a strong male bias in human ASDs (40) and many ASD animal models bear sexually dimorphic behavioral abnormalities (41, 42), we further tested whether female adult Mdga1-cKO mice displayed similar autistic-like behavioral phenotypes (Supplemental Fig. 14).

Indeed, female adult Mdga1-cKO mice exhibited normal sociability, USV numbers, USV syllable compositions, sensorimotor gating, and novel object-recognition memory (Supplemental Fig. 14). Also, unlike their male counterparts, female adult Mdga1-cKO mice showed reduced anxiety-like behavior (Supplemental Fig. 14D and 14E). These results suggest that MDGA1 could be a potential sex-specific biomarker for ASDs.

## Discussion

Here we propose that MDGA1, previously characterized as a GABAergic synapse-specific suppressor, may serve as a translational biomarker for MDGA1-associated neurodevelopmental and/or neuropsychiatric disorders, including ASDs. Given that various components of MDGA1- containing synaptic complexes have consistently been linked to ASDs (43), we examined the possibility that MDGA1 might also be implicated in ASDs. The current study provides new insights into the pathways by which two ASD-associated MDGA1 variants induce previously recognized pathogenesis, including abnormal neuronal migration and imbalanced excitation/inhibition (E/I) ratios (34, 44–49). Intriguingly, both MDGA1 variants manifested distinct loss-of-function and gain-of-function phenotypes during various stages of postnatal development in mice. These results differentiate MDGA1 from other proteins encoded by known ASD risk genes (50), in that MDGA1 dysfunction impinges on a spectrum of neurodevelopmental processes. The pathophysiological mechanisms underlying ASD-related alterations at glutamatergic synapses have been comprehensively investigated (46, 51–53). Decreased GABA levels and loss of PV^+^ interneurons, which are known ASD pathologies related to GABA system dysfunction, can increase the E/I ratio and network hyperactivity (54, 55). Notably, a number of synaptic genes implicated in defective GABA systems also affect the functions of glutamatergic synapses(55), making it difficult to precisely evaluate the significance of GABAergic synapse dysfunction in ASD pathophysiology. The MDGA1 variants identified and analyzed in this study did not directly alter the levels of GABA_A_ receptors, numbers of PV^+^ interneurons, or activities of neurons (see also accompanying paper). These variants also had no effect on the binding of MDGA1 to its ligands, Nlgn2 and APP, both of which are also encoded by ASD risk genes (56, 57). Our results provide a novel mechanistic basis for the malfunction of GABAergic synapse components in ASDs.

Both MDGA1 variants induced abnormal patterns of ultrasonic communication in pups, similar to the communicative deficits observed in children with autism. In utero overexpression of MDGA1 V116M/A688V impaired USVs in pups and induced abnormal neuronal migration, leading us to speculate that there may be a positive correlation between the two processes. In contrast, in utero overexpression of MDGA1 Y635C/E756Q failed to alter pup USVs or neuronal migration. In view of these results, we tentatively conclude that the two MDGA1 mutations induce distinct abnormal phenotypes in the early postnatal phase of mouse development.

The two MDGA1 variants exerted distinct effects on synaptic properties. MDGA1 WT and MDGA1 V116M/A688V suppressed GABAergic synapses, whereas MDGA1 Y635C/E756Q did not exhibit this activity. Strikingly, MDGA1 V116M/A688V impaired the dendritic spine structure and excitatory synaptic transmission. Whereas forebrain-specific Mdga1-cKO mice unambiguously demonstrated effects only in GABAergic synapses (33), MDGA1 V116M/A688V had unexpected effects in excitatory synapses. Previous studies established that glutamate plays an important role in neuronal migration, and neuron migration defects are linked to disruptions in glutamatergic neurotransmission (51, 58, 59). MDGA1 forms complexes with the gap junction subunit, connexin 43 (16), to provide a neurogenic niche within subventricular zones during cortical neuron migration, and contribute to sustaining glutamatergic synaptic activity (60). Thus, it is plausible that during the early phase of CNS development, MDGA1-mediated biological processes might be driven by glutamate.

Alternatively, a small population of endogenous MDGA1 expressed at glutamatergic synapses could possibly contribute to this process. Further investigation is warranted to unravel the underlying mechanisms. In the MDGA1 Y635C/E756Q mutant, E756Q contributes to destabilizing the triangular extracellular structure of MDGA1 (as suggested by in silico prediction and confirmed by TEM analyses). Our findings thus support the recent claim that maintenance of the compact three-dimensional structure of MDGA1 is physiologically significant and required for its function at the synaptic cleft in mature neurons (61). Although our approach of introducing two MDGA1 missense variants into a single expression vector provides insights into how these variants dysregulate MDGA1 functions, we admit that this strategy is unable to entirely replicate the biological complexity of a compound heterozygous state in patients with MDGA1 variants, where each mutation occurs on a separate allele.

Adult male Mdga1-cKO mice exhibited a range of autistic-like behavioral abnormalities. These changes included mild deficits in sociability, reduced USV call numbers and USV duration, and abnormal sensorimotor gating, supporting a genetic association of MDGA1 with ASDs. The behavioral profiles of adult male Mdga1-cKO mice differed from those recently reported for Mdga2- KO mice (62), which is consistent with the report that MDGA1 and MDGA2 play distinct roles in negatively regulating GABAergic and glutamatergic synapses, respectively (33). However, given that MDGA2 forms complexes with Nlgn1 (or Nlgn3) and Nrxns, and Nlgn3 is closely linked to ASDs, it is highly likely that MDGA2 is also associated with ASDs. Intriguingly, the total level of MDGA paralogs appears to be critical for maintaining synaptic homeostasis in neurons (33). Thus, the mechanisms by which the unique action of each MDGA paralog contributes to synaptic homeostasis in neurons and whether its impairment triggers ASDs require further elucidation.

The behavioral impairments we herein documented in forebrain excitatory neuron-specific Mdga1-cKO mice support the pathogenicity of MDGA1 deficiency. However, it is challenging to directly translate these animal model-derived results to human patients, as the same genetic mutation can often result in different clinical presentations. The functional consequences of MDGA1 mutations may depend on variable factors, such as protein stability, interacting protein network and/or neuronal functions. Given the critical role of MDGA1 in suppressing GABAergic synaptic properties, it is possible that even slight differences in MDGA1 dysfunction could contribute to variations in disease severity. While our findings established a strong connection between biallelic MDGA1 mutations and neurodevelopmental processes, further study is required to fully understand how different variant combinations influence clinical outcomes. In this regard, patient-derived neuronal models will be crucial for unraveling the genotype-phenotype relationship in MDGA1-associated neurodevelopmental disorders. In view of this, the immediate goals of future studies should be to introduce idiopathic ASD-associated MDGA1 mutation(s) into induced neuronal cells and determine whether this can replicate major cellular disease phenotypes in human neurons. Although further explorations are warranted, our study provides important clues into how proper control of GABAergic synaptic inhibition is mechanistically linked to ASD pathogenesis. Intriguingly, adult female Mdga1-cKO mice did not show any of the prominent behavioral deficits seen in their male counterparts. Together with our extensive analyses of male and female Mdga1^Y636C/E751Q^ knock-in mice (see accompanying paper for details), these results suggest that our MDGA1 mouse models could be used in the future to elucidate the sexually dimorphic nature of ASDs.

## Methods

### Identification of cases with MDGA1 variants

505 cases of trio exome sequencing performed in our department since 2014 were reviewed in search for MDGA1 mutations in homozygosity or compound heterozygosity state, according to the haploinsufficiency of this gene. All studies were performed on patients with neurodevelopmental disorders of probable genetic origin. Two unrelated cases with compound heterozygous MDGA1 mutations were identified.

### Genetic analysis

Whole exome sequencing was performed using genomic DNA isolated from whole blood from proband and parents. Genomic DNA extraction was carried out from blood using the Magna Pure 24 equipment (Roche Diagnostics). Quantity of extracted gDNA was measured with a fluorimeter (Quibit 3.0). The absorbance ratios at 260/280 and 260/230 were also studied to determine the quality of the DNA obtained, using NanoDrop ND-2000 equipment. In addition, Integrity of the genomic DNA was analyzed by electrophoresis in 0.8% agarose gels. Libraries were prepared using KAPA Hyper plus Kit (Roche Diagnostics) following the manufacturer’s specifications and capture enrichment protocol with specific probes (KAPA HyperExome; Roche Diagnostics). Then, we perform subsequent massive parallel sequencing in a NextSeq550 equipment (Illumina). Signal processing, base calling, alignment, and variant calling were performed with Genologica variant analysis software (GenoSystem). This software developed by Genologica contains an optimized algorithm that includes (among other steps), the following: (a) Initial quality control of the sequences; (b) Filtering the sequences by eliminating indeterminacies, adapters and low-quality areas; (c) Second quality control of the sequences; (d) Mapping on the Hg19 reference genome; (e) Obtaining variants and CNVs; (f) Mapping coverage study; and (g) Annotating variants. Finally, variant prioritization was based on stringent assessments at both the gene and variant levels and taking into consideration patient’s phenotype and the associated inheritance pattern. Candidate variants were visualized using IGV (Integrative Genomics Viewer). Candidate variants were evaluated based on stringent assessments at both the gene and variant levels, taking into consideration both the patient’s phenotype and the inheritance pattern. Variants were classified following the guidelines of the American College of Medical Genetics and Genomics (ACMG). A board of molecular clinical geneticist evaluated each variant classified as pathogenic, likely pathogenic, or a variant of uncertain significance, and decided which, if any, had to be reported. In every case, causal variants were discussed with the referring physician and/or clinical geneticist. Effects of MDGA1 missense variants were predicted using various in silico tools, including SIFT (<u>S</u>orting Intolerant from <u>T</u>olerant; https://sift.bii.a-star.edu.sg); FATHMM (<u>F</u>unctional <u>A</u>nalysis through <u>H</u>idden <u>M</u>arkov <u>M</u>odels; https://fathmm.biocompute.org.uk); Eigen (https://www.columbia.edu/eigen); LRT (<u>L</u>ikelihood <u>R</u>atio <u>T</u>est) (63); MutationTaster (https://www.mutationtaster.org); and MVP (<u>M</u>issense <u>V</u>ariant <u>P</u>athogenicity prediction) (64).

### Neuroimaging

The brain MRIs entail different sequences, including DTI and 3D-SPGRT1, regardless of the reason for the study. DTI images were obtained with a 3T system (GE Medical System, Milwaukee, Wisconsin) by using a SS-SE echoplanar Diffusion weighted image (DWI) sequence (TR:12000; FOV: 240 mm; sections thickness: 3 mm, 0 spacing; matrix 128 × 128; bandwidth: 250; 1 nex; diffusion encoding in 45 directions) with maximum b = 1000 sec/mm^2^. 3D-tractography was performed in an off-line workstation by using commercially available processing software as provided by the manufacturer (Functool 3D Fiber Tracking, GE, France) based on fiber assignment by contiguous tracking (FACT) method, achieved by connecting voxel to voxel. The threshold values were 0.3 for FA and 45° for the trajectory angles, between the regions of interest (ROIs). DTI tracts were also co-registered to the 3D-T1 weighted data set. MRI evaluation and tractography analysis was conducted by a radiologist specializing in these techniques who was unaware of the patient’s genetic diagnosis.

### Ethical procedure

The study was carried out in accordance with the Declaration of Helsinki of the World Medical Association and approved by the Local Ethics Committees (Comunidad de Madrid, Spain; Ref. 30062019). Informed consent was obtained from each family after full explanation of the procedures.

### Animals

All mice were maintained and handled in accordance with protocols (DGIST-IACUC- 23112809-0003) approved by the Institutional Animal Care and Use Committee of DGIST under standard, temperature-controlled laboratory conditions. Mice were kept on a 12:12-h light/dark cycle (lights on at 7:00 am) and received water and food ad libitum. All experimental procedures were performed on male mice, using littermate control without Cre expression. Conditional Mdga1-KO mice was previously described (16, 20). Pregnant rats (Daehan Biolink) were used to prepare in vitro cultures of dissociated hippocampal neurons.

### Identification of MDGA1 in a single–cell human brain atlas

We utilized the single–cell brain atlas from our previous study (24), integrated various single–cell transcriptomic datasets, and compared the normalized gene counts for MDGA1. Among the collected data, cells and cell clusters annotated as poor quality in the original studies and those lacking sample information were excluded. Metadata, including sample ID, batch, platform, demographics, brain region, and postmortem interval were standardized with age information categorized into developmental stages. Gene names were standardized with respect to the NCBI and HGNC databases, and doublets detected by Scrublet version 1.8.2 (https://github.com/swolock/scrublet) were excluded. Normalization, log transformation, and selection of highly variable genes were executed using Scanpy version 1.8.2 (https://scanpy.readthedocs.io). Integration with batch effect correction was conducted using scvi-tools version 1.0.3 (https://scvi-tools.org), and clustering was conducted with the Leiden algorithm (resolution = 0.6). Primary cell types were annotated based on the expression of canonical cell type maker genes defined by the Allen Brain Institute (65). Genes that were differentially expressed in each cluster were identified using Wilcoxon rank sum test and cluster-specific genes were defined as those with a False Discovery Rate < 0.05 and a log2 fold change > 0.2.

### Gene set enrichment test

Gene set enrichment tests were performed for risk genes associated with various neurological disorders. ASD risk genes selected from multiple comparisons were retrieved from Fu et al. (n = 184) (66) and Satterstrom et al. (n = 96) (26) and categorized as being involved with gene expression regulation (n = 54) and neuronal communication (n = 24) as previously defined (26). Alzheimer’s disease risk genes were obtained from Lambert et al. (n = 32) (67). Epilepsy risk genes were identified from Heyne et al. (n = 50) (68). Schizophrenia gene set was retrieved from Singh et al. (n = 32) (69). Genes significantly enriched for PTVs or damaging missense variants (p value < 0.01) in cases and listed as bipolar disorder risk genes in the BipEx dataset (https://bipex.broadinstitute.org; n = 58) were selected. The developmental disorder gene set (n = 372) was sourced from Fu et al. (66). Intellectual disability risk genes (n = 28) were collected from Willsey et al. (70). ADHD risk genes (n = 74) were gathered from Demontis et al. (71). Gene identifiers for these gene sets were matched between gene sets by converting each gene alias into the corresponding HGNC gene symbol. For the enrichment test, Fisher’s exact test (one-sided) with multiple comparison by Bonferroni procedure was applied.

### Expression vectors

pDisplay-MDGA1 V116M/A688V, pDisplay-MDGA1 Y635C/E756Q, pDisplay- MDGA1 V116M, pDisplay-MDGA1 A688V, pDisplay-MDGA1 Y635C, and pDisplay-MDGA1 E756Q were generated via site-directed mutagenesis (Stratagene) using pDisplay-MDGA1 WT plasmid(18) as a template. L313-MDGA1 V116M/A688V and L313-MDGA1 Y635C/E756Q were similarly generated using the L313-MDGA1 WT construct (18) as a template. pDisplay-MDGA2 Y631C/E751Q was generated via site-directed mutagenesis using pDisplay-MDGA2 WT plasmid (18) as a template. pAAV-MDGA1 V116M/A688V and pAAV-MDGA1 Y635C/E756Q were similarly generated using pAAV-MDGA1 WT plasmid (20) as a template. pCAGG-His-HA-MDGA1 variants (WT, V116M/A688V and Y635C/E756Q) were constructed by amplification of the indicated full-length sequences via polymerase chain reaction (PCR), followed by digestion with EcoRI and NotI and subcloning into the pCAGGS-His-HA vector (Addgene). pCIG2-HA-MDGA1 variants (WT, V116M/A688V and Y635C/E756Q) were constructed by amplification of the indicated full-length sequences via PCR, followed by digestion with EcoRI and XmaI, and subcloning into the pCIG2 vector (Addgene). The pCMV-IgC APP (20) and pCMV-IgC Nlgn2 (18) constructs have been described previously.

### Antibodies

The following commercially available antibodies were used: mouse monoclonal anti-APP (clone 22C11; Millipore; Cat# MAB348; RRID: AB_94882); mouse monoclonal anti-β-actin (clone AC-74; Sigma-Aldrich; Cat# sc-47778; RRID: AB_476743); mouse monoclonal anti-Nlgn1 (Synaptic Systems; Cat# 129 111; RRID: AB_887747); rabbit polyclonal anti-Nlgn2 (Synaptic Systems; Cat# 129 202; RRID: AB_993011); guinea pig polyclonal anti-VGLUT1 (Millipore; Cat# ab5905; RRID: AB_2301751); mouse monoclonal anti-gephyrin (Synaptic Systems; Cat# 147 011; RRID: AB_887719); rabbit polyclonal anti-VGAT (Synaptic Systems; Cat# 131 003; RRID: AB_887869); rabbit polyclonal anti-GABA_A_ receptor γ2 (Synaptic Systems; Cat# 224 003; RRID: AB_2263066); mouse monoclonal anti-GAD67 (clone 1G10.2; Millipore; Cat# MAB5406; RRID: AB_2278725); mouse monoclonal anti- NMDAR1 (Clone 54.1; Millipore; Cat# MAB363; RRID: AB_94946); rabbit polyclonal anti-PSD-95 (goat polyclonal anti-EGFP (Rockland; Cat# 600-101-215; RRID: AB_218182); chicken polyclonal anti-EGFP (Aves Labs; Cat# GFP-1020; RRID: AB_10000240); mouse monoclonal anti-HA (clone 16B12; BioLegend; Cat# 901501; RRID: AB_2565006); rabbit monoclonal anti-HA (clone C29F4; Cell Signaling Technology; Cat# 3724; RRID: AB_1549585); rat monoclonal anti-somatostatin (clone YC7; Millipore; Cat# MAB354; RRID: AB_2255365); mouse monoclonal anti-PV (clone PV235; Swant; Cat# 235; RRID: AB_10000343); rabbit monoclonal anti-c-Fos (clone 9F6; Cell Signaling Technology; Cat# 14609; RRID: AB_2798537); rabbit polyclonal anti-phosphoserine (Millipore; Cat# AB1603; RRID: AB_390205); mouse monoclonal anti-Synapsin I (Synaptic Systems; Cat# 106 011; RRID: AB_2619722); mouse monoclonal anti-Synapsin II (Synaptic Systems; Cat# 106 211; RRID: AB_2619774); rabbit polyclonal anti-phospho-Synapsin II (Ser425) (Thermo Fisher Scientific; Cat# PA5-64855; RRID: AB_2663626); mouse monoclonal anti-Calbindin (Swant; Cat# 300; RRID: AB_10000347); rabbit polyclonal anti-Wfs1 (Proteintech; Cat# 11558-1-AP; RRID: AB_2216046); chicken polyclonal anti-Tbr1 (Millipore; Cat# AB2261; RRID: AB_10615497); rabbit polyclonal anti-Brn2 (Abcam; Cat# ab94977; RRID: AB_10615497); and Cy3-donkey anti-human IgG antibodies (Jackson ImmunoResearch; Cat# 709-165- 149; RRID: AB_2340535). The following antibody was previously described: rabbit polyclonal anti-pan-SHANK (1172; RRID: AB_2810261) and rabbit polyclonal anti-PSD-95 (JK016; RRID: AB_2722693) (72).

### Cell culture

HEK293T cells were cultured in Dulbecco’s Modified Eagle’s Medium (DMEM; Welgene) supplemented with 10% fetal bovine serum (FBS; Tissue Culture Biologicals) and 1% penicillin-streptomycin (Thermo Fisher) at 37°C in a humidified 5% CO_2_ atmosphere. All procedures were performed according to the guidelines and protocols for rodent experimentation approved by the Institutional Animal Care and Use Committee of Daegu Gyeongbuk Institute of Science and Technology (DGIST).

### Cell-surface binding assays

Ig-fusion proteins of Nlgn2, APP, and IgC alone (control) were produced in HEK293T cells. Soluble Ig-fused proteins were purified using protein A-Sepharose beads (GE Healthcare) as previously described. Bound proteins were eluted with 0.1 M glycine (pH 2.5) and immediately neutralized with 1 M Tris-HCl (pH 8.0). Transfected HEK293T cells expressing the indicated plasmids were incubated with 10 μg/ml Ig-fused proteins for 2 h at 37°C with gentle agitation. Images were acquired using a confocal microscope (LSM800; Carl Zeiss).

### Staining for surface/intracellular protein levels

HEK293T cells were transfected with expression vectors for HA-MDGA1 WT or the indicated variants. After 48 h, cells were washed twice with PBS, fixed with 3.7% formaldehyde for 10 min at 4 °C, and blocked with 3% horse serum/0.1% bovine serum albumin (BSA; crystalline grade) in PBS for 15 min at room temperature. Surface-expressed protein was detected by staining with mouse anti-HA antibody at room temperature. After 90 min, cells were washed twice with PBS and incubated with Cy3-conjugated anti-mouse antibodies for 1 h at room temperature. Next, cells were permeabilized with PBS containing 0.2% Triton X-100 for 10 min at 4 °C and incubated with rabbit anti-HA antibody for 90 min at room temperature to label intracellularly expressed proteins, followed by FITC-conjugated anti-rabbit secondary antibodies. Images were acquired using a confocal microscope (LSM800; Carl Zeiss).

### Production of recombinant lentiviruses

Lentiviruses were produced by transfecting HEK293T cells with three plasmids–lentivirus vectors, psPAX2, and pMD2.G–at a 2:2:1 ratio. After 72 h, lentiviruses were harvested by collecting the media from the HEK293T cells and briefly centrifuging at 1,000 × g to remove cellular debris. Filtered media containing 5% sucrose were centrifuged at 117,969 × g for 2 h; supernatants were then removed, and the virus pellet was washed with ice-cold PBS and resuspended in 80 μl PBS, as previously described (18).

### Heterologous synapse formation analyses

HEK293T cells were transfected with EGFP (control) or cotransfected with pDisplay-HA-Nlgn2, alone or together with the indicated MDGA1 variants using PEI. After 48 h, the transfected HEK293T cells were trypsinized, seeded onto day in vitro 12 (DIV12) hippocampal neurons, co-cultured for an additional 24 h, and double-immunostained on DIV13 with antibodies against EGFP, HA, and synapsin. All images were acquired using a confocal microscope (LSM800; Zeiss). For quantification, the contours of transfected HEK293T cells were chosen as the region of interest (ROI). Fluorescence intensities of synaptic markers in each ROIs were quantified for both red and green channels using MetaMorph software (Molecular Devices). Normalized synapse density on transfected HEK293T cells was expressed as the ratio of red to green fluorescence.

### Transfection, immunocytochemistry, confocal microscopy imaging, and analyses

Hippocampal cultured neurons were prepared from E18 rat brains cultured on coverslips coated with poly-L-lysine, and grown in Neurobasal medium supplemented with B-27 (ThermoFisher Scientific), 0.5% FBS, 0.5 mM GlutaMax (ThermoFisher Scientific), and sodium pyruvate (ThermoFisher Scientific), as previously described (73). Cultured neurons were cotransfected with EGFP alone with indicated MDGA1 variant expressing plasmids using a CalPhos Transfection Kit (Takara) at DIV5 and immunostained at DIV14. For immunocytochemistry, cultured neurons were fixed with 4% paraformaldehyde/4% sucrose, permeabilized with 0.2% Triton X-100 in phosphate buffered saline (PBS), immunostained with the indicated primary antibodies, and detected with the indicated Cy3-, fluorescein isothiocyanate (FITC)-, or Alexa Fluor 647-conjugated secondary antibodies (Jackson ImmunoResearch). Transfected neurons were chosen randomly, and images were acquired using a confocal microscope (LSM800; Zeiss) with a 63 × objective lens; all image settings were kept constant. Z stack images were converted to maximal intensity projection and analyzed to obtain the size, intensity, and density of puncta immunoreactivities derived from marker proteins. Quantification was performed in a blinded manner using MetaMorph software.

### Immunohistochemistry, confocal microscopy imaging, and analyses

P60 mice were anaesthetized and immediately perfused, first with PBS for 3 min, and then with 4% paraformaldehyde for 5 min. Brains were dissected out, fixed in 4% paraformaldehyde overnight, and sliced into 30-um-thick coronal sections using a vibratome (Model VT1200S; Leica Biosystems).

Sections were permeabilized by incubating with 2% Triton X-100 in PBS containing 5% bovine serum albumen and 5% horse serum for 30 min. For immunostaining, sections were incubated for 8–12 h at 4°C with primary antibodies diluted in 0.1% Triton X-100 in PBS containing bovine and horse serum. The following primary antibodies were used: anti-VGLUT1 (1:300), anti-PSD-95 (1:300), anti-VGAT (1:300), anti-gephyrin (1:500), anti-PV (1:300), anti-SST (1:50), anti-c-Fos (1:400), anti-Brn2 (1:100), anti- Wfs1 (1:100), and anti-Calb (1:100). Sections were washed three times in PBS and incubated with appropriate Cy3- or FITC-conjugated secondary antibodies (Jackson ImmunoResearch) for 2 h at room temperature. After three washes with PBS, sections were mounted onto glass slides (Superfrost Plus; Fisher Scientific) with VECTASHIELD mounting medium (H-1200; Vector Laboratories). Images were acquired by confocal microscopy (LSM800; Zeiss). Synaptic puncta were quantified using MetaMorph software, and their density and average area were measured. For immunohistochemical analysis of electroporated mice (P21), the mice were heart-perfused with 4% paraformaldehyde. Brains were collected, embedded in 3% low melting-temperature agarose, and sectioned at 100 μm thickness with a vibratome (VT1200S; Leica). The sections were incubated with primary antibodies (anti-EGFP, 1:2000; anti-HA, 1:1000) in PBS containing 0.1% Triton X-100, 0.1% BSA, and 2.5% goat serum, and then with fluorescent secondary antibodies (Alexa 488 and Alexa 647). Slices were mounted in Antifade mounting medium with DAPI (VECTASHIELD) and images were acquired by confocal microscopy (A1R; Nikon).

### Preparation of adeno-associated viruses and titration

HEK293T cells were co-transfected with the indicated AAV vectors and pHelper and pAAV1.0 (serotype 2/9) vectors. 72 hours later, transfected HEK293T cells were collected, lysed, and mixed with 40% polyethylene glycol and 2.5 M NaCl, and centrifuged at 2,000 × g for 30 min. The cell pellets were resuspended in HEPES buffer (20 mM HEPES; 115 mM NaCl, 1.2 mM CaCl_2_, 1.2 mM MgCl_2_, 2.4 mM KH_2_PO_4_) and an equal volume of chloroform was added. The mixture was centrifuged at 400 × g for 5 min, and concentrated three times with a Centriprep centrifugal filter (Millipore) at 1,220 × g for 5 min each and with an Amicon Ultra centrifugal filter (Millipore) at 16,000 × g for 10 min. Before the titration of AAVs, contaminating plasmid DNA was eliminated by treating 1 μl of concentrated, sterile-filtered AAVs with 1 μl of DNase I (Sigma-Aldrich) for 30 min at 37°C. After treatment with 1 μl of stop solution (50 mM ethylenediaminetetraacetic acid) for 10 min at 65°C, 10 μg of protease K (Sigma-Aldrich) was added and AAVs were incubated for 1 h at 50°C. Reactions were inactivated by incubating samples for 20 min at 95°C. The final virus titer was quantified by qRT-PCR detection of EGFP sequences and subsequent reference to a standard curve generated using the pAAV-T2A-EGFP plasmid. All plasmids were purified using a Plasmid Maxi Kit (Qiagen GmbH).

### Preparation and transfection of organotypic hippocampal and prefrontal cortical slice cultures

Organotypic slice cultures from mouse hippocampus or prefrontal cortex were prepared from postnatal day 2 (P2)–P3 mice, as described previously (20, 74), in accordance with the guidelines of the Institutional Animal Care and Use Committee at the University of Colorado Anschutz Medical Campus (protocol number: 721; IBC protocol number: 1305). Mice were acquired from Jackson Laboratory (C57BL/6N wild type). Slices were transfected with pDisplay MDGA1 WT, pDisplay MDGA1 V116M/A688V or pDisplay MDGA1 Y635C/E756Q 13–20 days prior to two-photon imaging using biolistic gene transfer (180 psi). A total of 5 μg tdTomato and 5 μg GFP-gephyrin-intrabody or 20 μg of the indicated pDisplay MDGA1 vector were coated onto 6–7 mg gold particles. The age of the culture was expressed as equivalent postnatal (EP) day; postnatal day at slice culturing + days in vitro.

### Two-photon imaging

Imaging was performed at EP 18–24 on transfected hippocampal CA1 pyramidal neurons or mPFC layer II/III pyramidal neurons within 40 μm of the slice surface at 30°C in recirculating artificial CSF (ACSF) (127 mM NaCl, 25 mM NaHCO_3_, 1.25 mM NaH_2_PO_4_, 2.5 mM KCl, 25 mM D-glucose) aerated with 95% O_2_/5% CO_2_. For each neuron, image stacks (512 × 512 pixels; 0.047 μm / pixel) with 1 μm z-steps were acquired from one segment of secondary or tertiary apical and/or basal dendrites using a two-photon microscope (Bruker Nano, Inc) with a pulsed Ti::sapphire laser (MaiTai HP; Spectra Physics) tuned to 920 nm (4-5 mW at the sample). All images are maximal projections of three-dimensional image stacks after applying a median filter (2 × 2) to the raw image data.

### Quantification of dendritic spines

All distinct protrusions emanating from the dendritic shaft, regardless of shape, were counted and measured on red fluorescent images using ImageJ (NIH). In all neurons included for analysis, overall spine density did not significantly change over the imaging session. This ensured that the density of synapses under examination was not influenced by cell health (which was always monitored), as widespread spine/filopodia-like structures can form in cases where cell health is compromised. Estimated spine volume was measured from bleedthrough– corrected and background-subtracted red fluorescence intensities using the integrated pixel intensity of a boxed region of interest (ROI) surrounding the spine head, as described previously (20).

### Quantification of gephyrin puncta

Gephyrin fluorescence intensity was calculated from bleedthrough-corrected and background-subtracted green (GFP-gephyrin intrabody) fluorescence intensities using the integrated pixel intensity of a boxed region surrounding a GFP-gephyrin punctum as described previously (20). Gephyrin enrichment in dendritic shafts was calculated by normalizing GFP-gephyrin fluorescence intensities (as described above) for each punctum to the mean GFP fluorescence intensity determined from four background ROIs on the same dendritic shaft. GFP-gephyrin enrichment was defined as a gephyrin punctum in cases where the ratio of green fluorescence from a punctum to green fluorescence from the dendritic background (Gp/Gd) was > 1. Among these GFP puncta, those with green fluorescence intensities more than two standard deviations (2 SDs) greater than the local background green fluorescence levels measured from two ROIs near the puncta were classified as true gephyrin puncta and counted in images from the green channel using ImageJ (NIH). A summary of the criteria is as follows: gephyrin puncta, Gp/Gd > 1 and expression level > 2 SDs of background levels; no puncta, Gp/Gd < 1 or expression level < 2 SDs of the background level.

### Stereotactic surgery and virus injections

Adult (P30) mice were anesthetized by intraperitoneal injection of a saline-based 2% Avertin solution (2,2,2-tribromoethyl alcohol dissolved in tert- amylalcohol [Sigma]), and their heads were fixed in a stereotactic apparatus. Recombinant AAV virus was injected into the hippocampal CA1 region (coordinates: AP -2.1 mm, ML ± 1.3 mm, and DV 1.8 mm) with a Hamilton syringe at a flow rate of 100 nl/min (injected volume, 300 nl) using a Nanoliter 2010 Injector (World Precision Instruments). Each injected mouse was restored to its home cage for 2- 4 weeks and used subsequently for immunohistochemical analyses, electrophysiological recordings, or behavioral analyses.

### Electrophysiology

#### 1. Cultured hippocampal neurons

Hippocampal cultured neurons from wild type rat embryos were transfected with MDGA1 and MDGA2 patient mutant variants, followed by analysis at DIV13–16, as previously described (75). Using a Model P-97 pipette puller, pipettes were pulled from borosilicate glass (i.d. 0.86 mm, o.d. 1.5 mm; Sutter Instruments). The resistance of patch pipettes containing an external solution ranged from 3 to 6 MΩ. The internal solution included 145 mM CsCl, 0.3 mM Na-GTP, 10 mM EGTA, 10 mM HEPES, 5 mM NaCl, and 4 mM Mg-ATP, with a pH of 7.2–7.4 adjusted with CsOH and an osmolarity of 290–295 mOsmol/L for mEPSCs and mIPSC. The external solution included 130 mM NaCl, 1 mM MgCl_2_, 2 mM CaCl_2_, 4 mM KCl, 10 mM D-glucose, and 10 mM HEPES with a pH of 7.2–7.4 calibrated with NaOH and an osmolarity of 300–305 mOsmol/L. The whole-cell arrangement was built at room temperature using M-TSC manipulators (SENSAPEX). Electrophysiological data were obtained using an Axon Multiclamp 700B amplifier and pCLAMP software and then digitized using an Axon DigiData 1550B data capture board (Axon Instruments); mEPSCs and mIPSCs were recorded at a holding potential of -70 mV, respectively.

Synaptic currents were analyzed offline using Clampfit 10.8 software (Molecular Devices). For the recordings of mEPSCs, the external solution contained 1 μM TTX and 50 μM picrotoxin to block the GABA_A_ receptor and Na^+^ currents, respectively. For the recordings of mIPSCs, 1 μM TTX, 10 μM CNQX, and 50 μM D-AP5 were included in the extracellular solution to block Na^+^ currents, AMPA receptors, and NMDA receptors, respectively. <u>2. Acute hippocampal CA1 slices</u>. Hippocampal slices (300 μm) were prepared from mice aged 10–12 days, 18–20 days, or 8–10 weeks. Following anesthesia with isoflurane, the mice were euthanized, and their brains were swiftly extracted and placed in a chilled solution with low calcium and high magnesium levels, which was oxygenated (95% O_2_ and 5% CO_2_). This solution consisted of 3.3 mM KCl, 1.3 mM NaH_2_PO_4_, 26 mM NaHCO_3_, 11 mM D-glucose, 0.5 mM CaCl_2_, 10 mM MgCl_2_, and 211 mM sucrose. Hippocampal slices were prepared using a vibratome (VT1000S; Leica) and moved to a storage chamber filled with oxygenated artificial cerebrospinal fluid (aCSF) containing 124 mM NaCl, 3.3 mM KCl, 1.3 mM NaH_2_PO_4_, 26 mM NaHCO_3_, 11 mM D-glucose, 2 mM CaCl_2_, and 1 mM MgCl_2_. Slices were incubated at 30°C for at least 60 minutes prior to experimentation. Subsequently, slices were moved into the recording chamber and subjected to constant perfusion with standard aCSF that was oxygenated with a mixture of 95% O_2_ and 5% CO_2_.

All experiments were performed at 32°C, and slices were used within 4 h. Only cells with access resistance (Ra) smaller than 30 MΩ were analyzed. All the recordings were conducted using a Multiclamp 700B amplifier and DigiData 1550B Digitizer. To assess miniature inhibitory postsynaptic currents (mIPSCs), whole cell recordings were performed from hippocampal CA1 pyramidal neurons using glass pipettes (3–5 MΩ) filled with a solution containing 145 mM CsCl, 5 mM NaCl, 10 mM HEPES, 10 mM EGTA, 4 mM Mg-ATP, and 0.3 mM Na-GTP, adjusted to pH 7.2–7.3 with CsOH. Cellular voltage was clamped at -70 mV. mIPSCs were isolated by inhibiting NMDAR, AMPAR, and Na^+^ channel using external application of 50 μM D-AP5, 10 μM CNQX, and 1 μM tetrodotoxin. For measurement of evoked inhibitory postsynaptic currents (eIPSCs) and delayed asynchronous IPSCs (aIPSCs), patch pipettes were filled with an internal solution consisting of 130 mM Cs-methanesulfonate, 5 mM TEA-Cl, 8 mM NaCl, 0.5 mM EGTA, 10 mM HEPES, 4 mM Mg-ATP, 0.4 mM Na-GTP, 1 mM QX-314, and 10 mM disodium phosphocreatine, adjusted to pH 7.2–7.3 with CsOH. Cells were voltage-clamped at 0 mV. Electrical stimulation was applied using a concentric bipolar electrode (FHC), placed on the stratum radiatum (SR) or stratum lacunosum moleculare (SLM) in the hippocampal CA1 to record somatic inhibition and dendritic inhibition respectively. eIPSCs and aIPSCs were isolated by inhibiting NMDAR, and AMPAR using external application of 50 μM D-AP5, and 10 μM CNQX. To record eIPSCs input-output (eIPSCs I-O), electrical stimulation was applied ranged 10 to 60 μA with 10 μA increments. Average eIPSCs I-O were measured from three consecutive sweeps. For assessing paired-pulse ratios (PPRs), pairs of stimuli were administered at 10 Hz frequency. PPRs were calculated as 2^nd^ eIPSC/1^st^ eIPSC. Average PPRs were analyzed from three consecutive sweeps. To assess delayed aIPSCs, 40 Hz train stimulation was applied. Synaptic events during 1 second area after train stimulation were quantified, normalized to the 1 second area before the train stimulation.

### Circular dichroism spectroscopy

Far-UV circular dichroism (CD) from 200 nm to 260 nm was performed on a spectrophotometer (J-1500; JASCO) with a temperature controller and 1 mm path- length quartz cuvette. Samples of wild-type and mutant MDGA1 (0.2 mg/ml) in PBS were prepared for CD measurements. Bandwidth was 5 nm and scanning speed was 20 nm/min with 4 seconds digital integration time. All spectra were measured by averaging 5 time-scanning with buffer subtractions and temperature was kept at 20°C. Millidegrees were converted to molar residue ellipticity.

### *In utero* electroporation

E15.5 timed-pregnant ICR mice were anesthetized with isoflurane and the uterine horns were exposed by laparotomy. Endotoxin-free plasmids (pCIG2, pCIG2-HA- MDGA1WT, pCIG2-HA-MDGA1 V116M/A688V, or pCIG2-HA-MDGA1 Y635C/E765Q; 2 μg/μl) and 0.5% Fast Green were injected into a lateral ventricle of E15.5 embryos. Electroporation was performed by placing the anode on the side of DNA injection and the cathode on the other side of the head to target cortical progenitors. Four pulses of 40 V for 50 ms with 500 ms interval were applied using an electroporator (ECM 830; BTX). After electroporation, the uterine horns were returned into maternal abdomen.

### Mouse behavioral tests

Male and female mice aged 7–9 weeks were used for all behavioral tests. Tests were conducted in a soundproofed room under dim lighting (< 5 lux). All behavioral examinations were conducted in a controlled environment and mice were acclimated to the experimental room prior to testing. The order of testing was randomized, and experiments conducted at consistent timeframes to minimize potential circadian influences.

<u>1. Open-field test</u>. Mice were placed in a white acrylic open-field box (40 × 40 × 40cm) and allowed to freely explore the environment for 10 min in dim light (< 5 lux). The distance traveled and time spent in the center zone by freely moving mice were recorded with a top-view infrared camera and analyzed using EthoVision XT 10.5 software (Noldus).
<u>2. Y-maze test.</u> A Y-shaped white acrylic maze with three 35 cm long arms at a 120’ angle from each other was used. Mice were introduced into the maze and allowed to explore freely for 8 min. A move was counted as an entry when all four limbs of the mouse were within the arm. Movement of mice was recorded with a top-view infrared camera and analyzed using EthoVision XT 10.5 software.
<u>3. Novel object-recognition test.</u> An open-field chamber was used for this experiment. Mice were habituated to the chamber for 10 min. For training sessions, two identical objects were placed in the center of the chamber at regular intervals and mice allowed to explore the objects for 10 min. After the training session, mice were returned to their home cages for 24 h. For novel object recognition tests, one of the two objects was replaced with a new object, which was placed in the same position in the chamber. Mice were returned to the chamber and allowed to explore freely for 10 min. Movement was recorded using an infrared camera and the number and duration of contacts analyzed using EthoVision XT 10.5. The discrimination index represents the difference between time spent exploring novel and familiar objects during the test phase.
<u>4. Elevated plus-maze test.</u> The white acrylic maze with two open arms (30 × 5 × 0.5 cm) and two closed arms (30 × 5 × 30 cm) was elevated by 75 cm over the floor. Mice were individually placed at the center of the elevated plus-maze and allowed to freely move for 5 min. All behaviors were recorded with a top-view infrared camera, and the time spent in each arm and number of arm entries analyzed using EthoVision XT 10.5.
<u>5. Light and dark box transition test.</u> The apparatus consisted of a roofless box divided into a closed dark chamber and a brightly illuminated chamber. A small entrance allowed free travel between the two chambers. The light chamber was illuminated at 350 lux. Movement of mice was recorded using a top-view infrared camera and EthoVision XT 10.5 applied for analysis of the time spent in each chamber and the number of transitions.
<u>6. Three-chamber test.</u> The testing apparatus consisted of a white acrylic box divided into three chambers (20 × 40 × 22 cm each) with small openings on the dividing walls. Wire cups were employed to enclose social conspecifics in the corners of both side chambers. Mice were placed in the central chamber for a 10 min habituation period. Following this time, an age-matched social conspecific was placed in the wire cup on the left side-chamber and sociability of the subjects assessed by measuring subject exploration times for the enclosed conspecific and empty cup during the second 10 min session. An exploration was counted when the subjects placed their nose in the vicinity of the wire cups. In the final 10 min session, a new social conspecific was placed into the empty wire cup on the right side-chamber and social recognition assessed by measuring subject exploration times of the familiar and novel conspecifics in the wire cup by the subject.
<u>7. Prepulse inhibition test.</u> Mice were individually acclimated to the experimental room for a minimum of 30 min under standard housing conditions with reduced extraneous noise. Each mouse was placed in a sound-attenuating chamber equipped with startle response measurement apparatus (SR-LAB- Startle Response System; SD Ins.), ensuring minimal movement during testing. On day 1, after a 5- min habituation period in the absence of stimuli, mice were exposed to a series of startle-eliciting stimuli (from 0 to 120 dB) to familiarize them with the startle response. On day 2, for the PPI trials, prepulse stimuli (73, 76, 79, or 82 dB) were presented before the startle-eliciting stimuli (120 dB). The order and inter-trial intervals of prepulse and startle stimuli were randomized, including control trials with no prepulse. Startle responses were recorded for each trial using PPI test software. Data analysis involved calculating the percentage PPI for each prepulse intensity using the formula: PPI (%) = (Rpulse−Rpre_−_pulse)/Rpulse × 100 (Rpulse is the average response magnitude in the absence of a prepulse and Rpre_−_pulse the average response magnitude in the presence of a prepulse).
<u>8. Ultrasonic vocalization test</u>. Following a 3-day period of single housing, each adult mouse was individually placed in a sound-attenuating chamber equipped with a microphone for USV recording. To introduce a stimulus for vocalization, a conspecific of the opposite sex, rendered anesthetized, was gently introduced into the chamber. The interactions and vocalizations of the focal mouse were recorded for a duration of 10 min. In experiments involving pups, sessions were conducted on postnatal days P3, P6, P9, and P12. Each pup was temporarily separated from the dam and placed in a cage with clean bedding on a heat pad maintained at 37 ^0^C to ensure body temperature regulation and its emitted ultrasonic vocalizations meticulously measured for 10 min. USV recordings were subjected to detailed analysis using DeepSqueak software (reference) for the detection and categorization of ultrasonic vocalizations. This software facilitated the extraction of key parameters, such as call frequency, duration, and type. Calls were systematically categorized based on their spectrotemporal features.
<u>9. Forced swim test.</u> Mice were individually placed in a glass cylinder (15ⅹ30 cm) containing water (24 ± 1 °C; depth, 15 cm). All mice were forced to swim for 6 min, and the duration of immobility measured during the final 4 min of the test. The latency to immobility from the start of the test (delay between the start of the test and appearance of the first bout of immobility, defined as a period of at least 1 s without any active escape behavior) and duration of immobility (defined as the time not spent actively exploring the cylinder or trying to escape from it) were measured. Immobility time was defined as the time the mouse spent floating in the water without struggling, making only minor movements that were strictly necessary to maintain its head above water.
<u>10. Tail-suspension test.</u> Mice were individually subjected to the tail suspension test, involving suspension by adhesive tape affixed approximately 1-2 cm from the tail tip. The testing apparatus consisted of a standardized setup and mice were allowed to hang freely. The total test duration was set at 5 min during which mice were examined for despair-related behaviors. The immobility time was recorded for the final 4 min of the test. Immobility was operationally defined as the absence of limb or body movement, and the recorded duration represented the time spent suspended without exhibiting active escape behaviors or exploratory movements. Additionally, latency to immobility was measured as the period during which the mouse did not engage in any active escape behavior for at least 1 second.
<u>11. Marble burying test.</u> Mice were individually acclimated to the experimental room for at least 30 min prior to testing. Standard mouse bedding was placed in home cages to provide a familiar environment. For the marble burying test, a 5 cm layer of fresh bedding was uniformly spread in the Plexiglas cage. Next, 15 glass marbles were evenly distributed on the bedding surface. Each mouse was introduced into the cage and allowed free exploration for a duration of 30 min. During the marble burying phase, the number of marbles buried (defined as being at least two-thirds covered by bedding) was counted and quantified.

### Negative staining

Human MDGA1 full ectodomain (16 μg/ml) and the Y635C/E756Q mutant (19 μg/ml) were applied to a glow-discharged carbon-coated 300 mesh copper grids (Electron Microscopy Science) and stained with 0.075% uranyl formate solution. Images were acquired using a Tecnai T12 Bio-TWIN transmission electron microscope equipped with an FEI Eagle 4_4K charge-coupled device camera and operating at 120 kV.

### Statistical analysis

No statistical method was used to pre-determine sample sizes. Rather, the sample sizes were selected based on previous studies published in the field (see Life Science Reporting Summary for references). Animals in the same litter were randomly assigned to the different treatment groups in the various experiments. All statistical analyses were performed using GraphPad Prism 7 software (RRID: SCR_002798). The normality of distributed data was determined using the Shapiro-Wilk normality test. Normally distributed data were compared using Student’s t- test or the analysis of variance (ANOVA) test, and non-normally distributed data by the Mann**–** Whitney U test, non-parametric ANOVA with Kruskal**–**Wallis test followed by post hoc Dunn’s multiple comparison test, or non-parametric ANOVA with post hoc Tukey’s multiple comparison test. If a single value made the data distribution non-normal and was found to be a significant outlier (p < 0.05) by Grubb’s test, it was regarded as an outlier.

## Data Availability

All data produced in the present study are available upon reasonable request to the authors

## Author contributions

Contributions: Conceptualization: A.F-.J and Jaewon K.; Data curation: Seungjoon K., H.K., J.P.P., S.A., G.J., Jinhu K., V.M.H., B.C.-P, H.J., J.L., Seoyeon K., M.J.d.l.P, A.-r.H, D.S.L. and S.J.; Funding acquisition: Seungjoon K, H.K., H.M.K., J.-Y.A., W.C.O., S.-K.K., J.W.U. and Jaewon K.; Investigation: Seungjoon K., H.K., G.J., Jinhu K., V.M.H., H.J., J.L., Seoyeon K., A.-r.H, and S.J.; Methodology: S.K., H.K., G.J., Jinhu K., V.M.C., B.C.-P, H.J., J.L., Seoyeon K., M.J.d.l.P, A.-r.H, D.S.L. and S.J.; Supervision: W.Y., H.M.K., J.Y.A., W.C.O., S.K.K., J.W.U., A.F-.J. and Jaewon K.; Writing – original draft: A.F-.J. and Jaewon K.; Writing – review & editing: Jaewon K.

## Acknowledgments

We thank Jinha Kim (DGIST) for technical assistance. This study was supported by National Creative Research Initiative Program of the Ministry of Science and ICT (RS-2022-NR070708 to Jaewon K.), the National Research Foundation of Korea (NRF) funded by the Korea Government (2022R1C1C200003499 to Seungjoon K.; RS-2024-00339642 to H.K.; NR-2020R1C1C1003426 to J.-Y.A.; RS-2023-00264980 to S.-K.K.; and 2023R1A2C2002535 to J.W.U.), Institute for Basic Science (IBS-R030- C1 to H.M.K.), and National Institutes of Health (R01MH124778, R21MH126073 and R21NS133681 to W.C.O.).

