## Supplemental Figures for "Autism-associated MDGA1 missense mutations impair distinct facets of central nervous system development"

**Supplemental figures and figure legends**

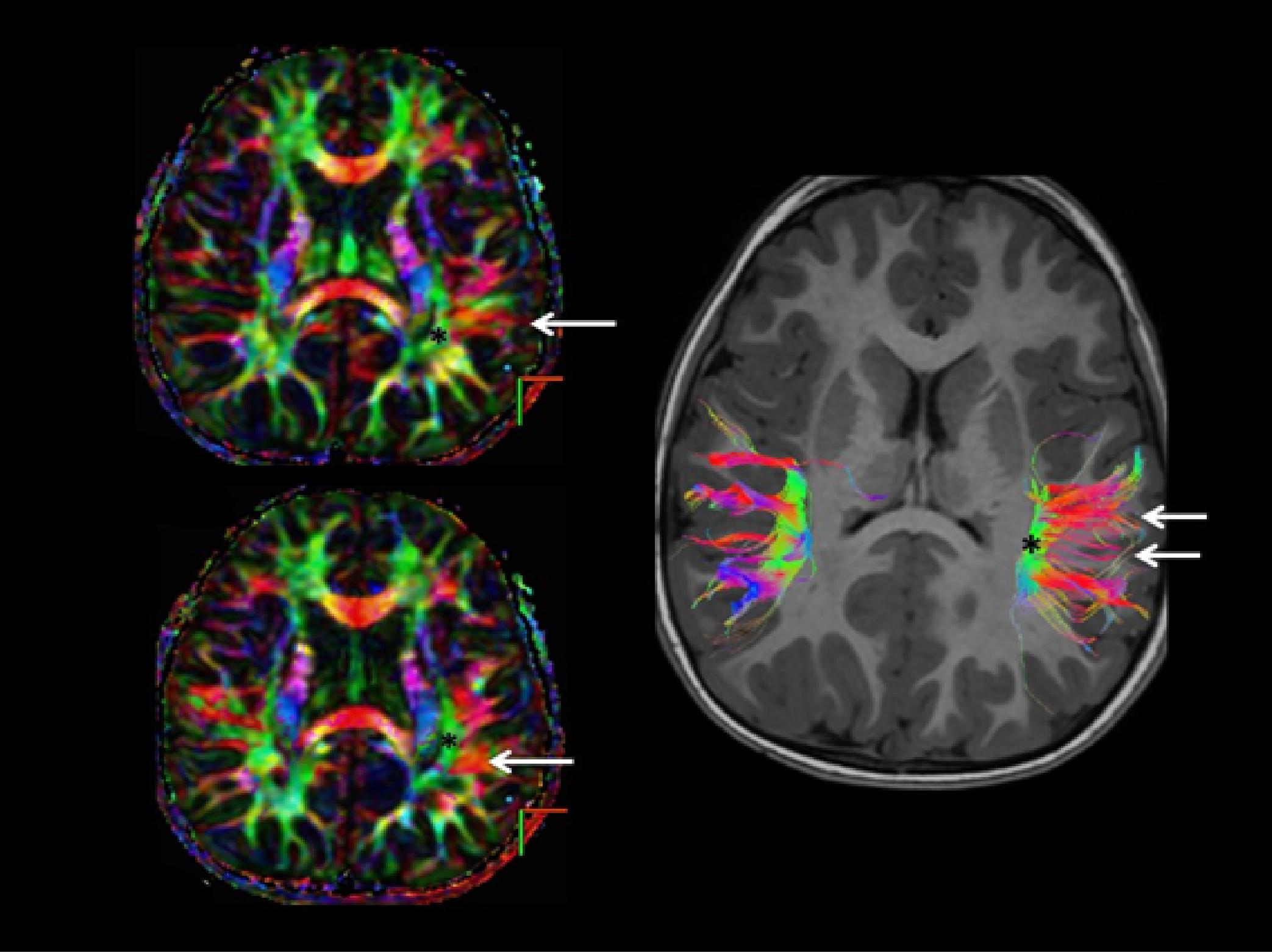

**Supplemental Figure 1. Diffusion tensor 2D-map and 3D-tractography reconstruction.**

Although a mild asymmetry of the anteroposterior component of the arcuate fascicle (asterisk) is visible on the 2D-fiber tracking image, the main finding is the clustering of the horizontal subcortical connections (white arrows) with the left inferior parietal lobe.

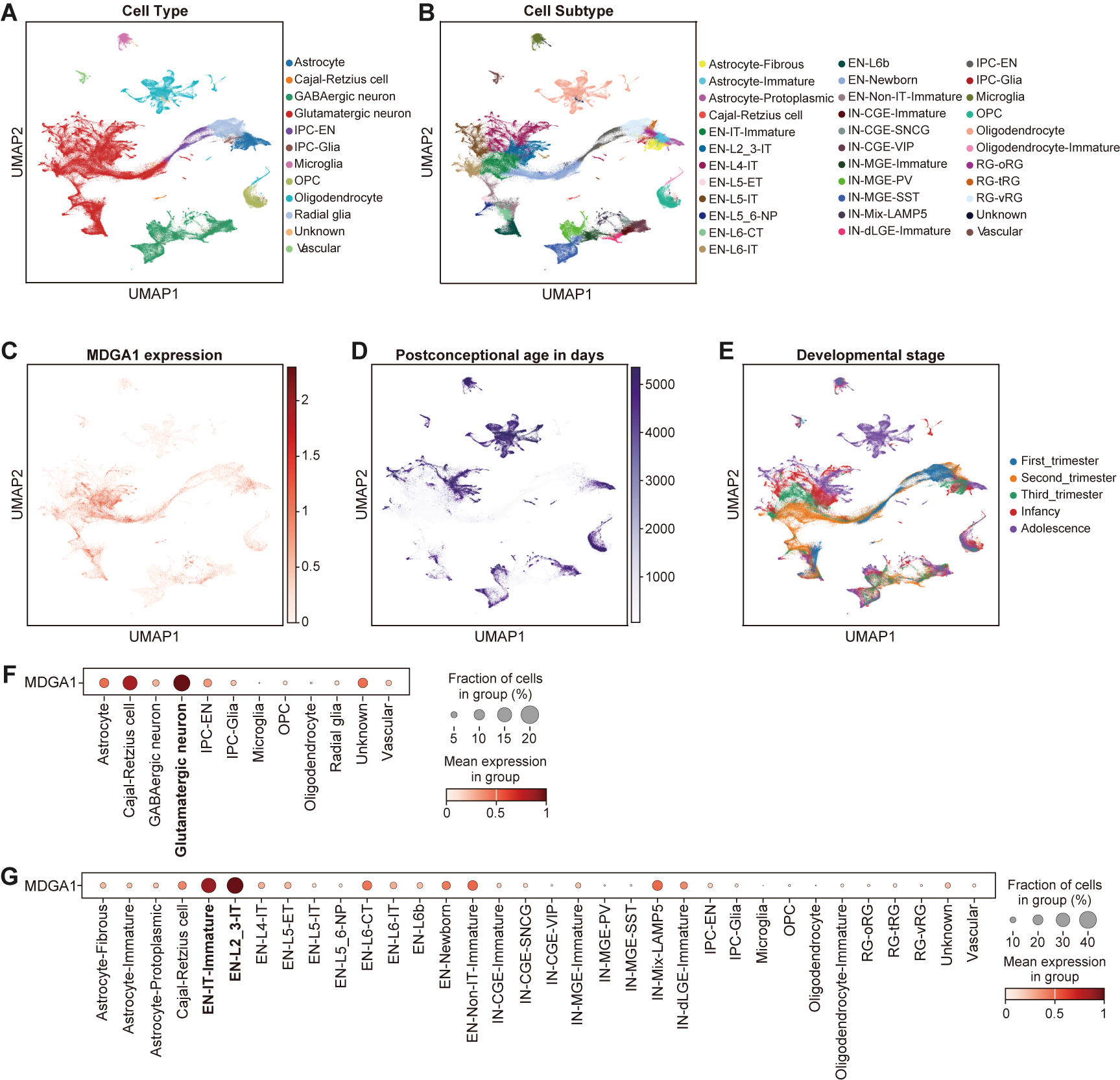

**Supplemental Figure 2. Analysis of MDGA1 expression in developing human neocortex.**

(**A–E**) UMAP visualization, with colors or labels indicating distinct clusters of major cell populations (**A**), more granular classification of neural cell subtypes (**B**), MDGA1 expression within each cell (**C**), postconceptional age of cells in days (**D**), and developmental stages of cells (**E**).

(**F and G**) Dot plot of MDGA1 expression across major cell types (**F**) and granular cell subtypes (**G**).

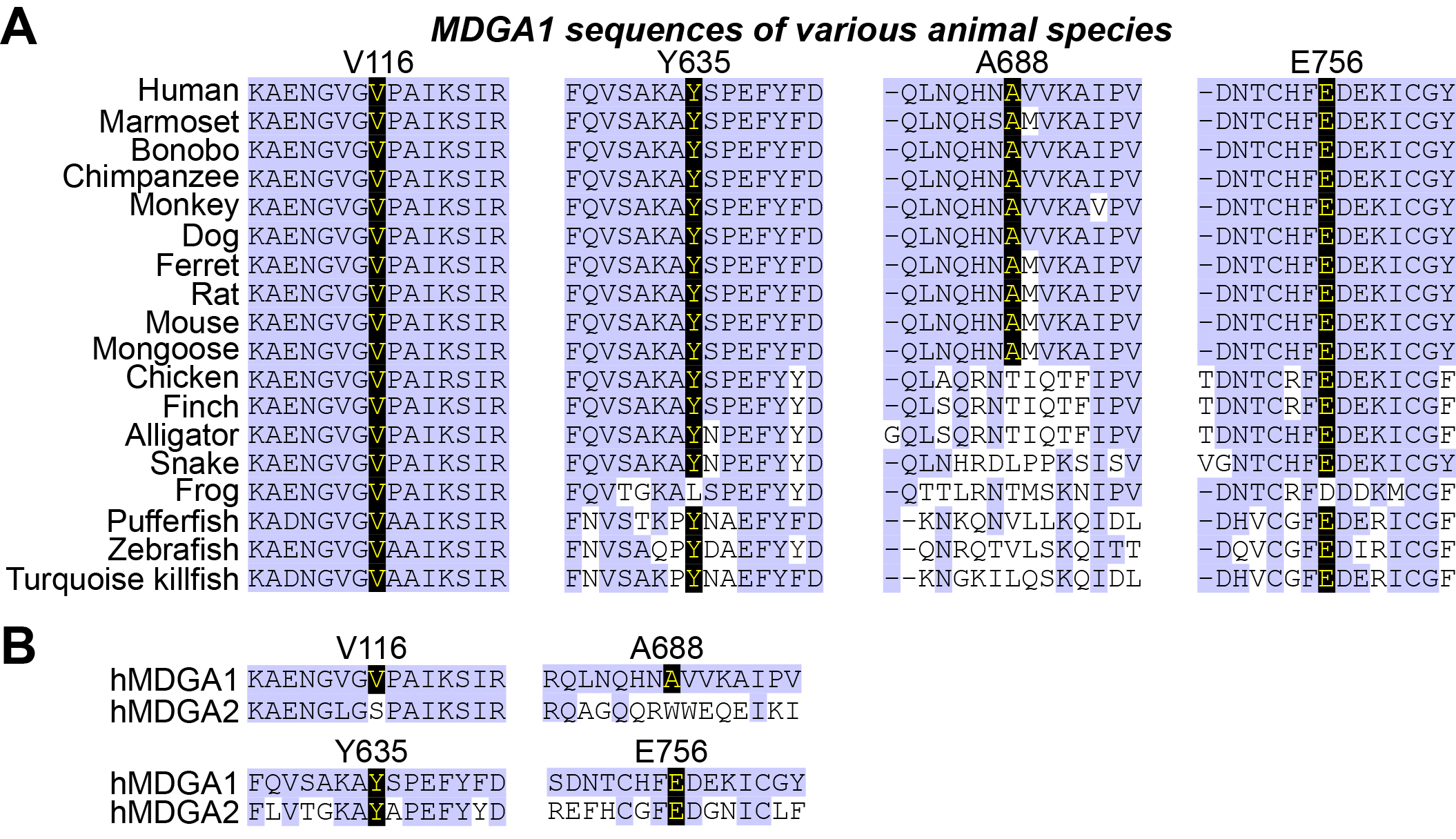

**Supplemental Figure 3. Sequence comparison and analysis of MDGA1 across different species.**

(**A**) Sequence alignment of MDGA1 from various species. The sequences are compared for the presence of conserved and variable regions. Specific amino acid residues of interest in this study (V116, Y653, A688 and E756) are highlighted across species, including human, mouse, rat, dog, zebrafish, chicken, alligator, marmoset, finch, ferret, turquoise killifish, bonobo, chimpanzee, snake, mongoose, pufferfish, monkey, and frog.

(**B**) Comparison of MDGA1 and MDGA2 sequences. The sequences are aligned to show conserved and divergent regions between the two MDGA family proteins, with specific attention given to the amino acids V116, Y653, A688 and E756. The comparison highlights similarities and differences that may contribute to the distinct functional roles of MDGA1 and MDGA2.

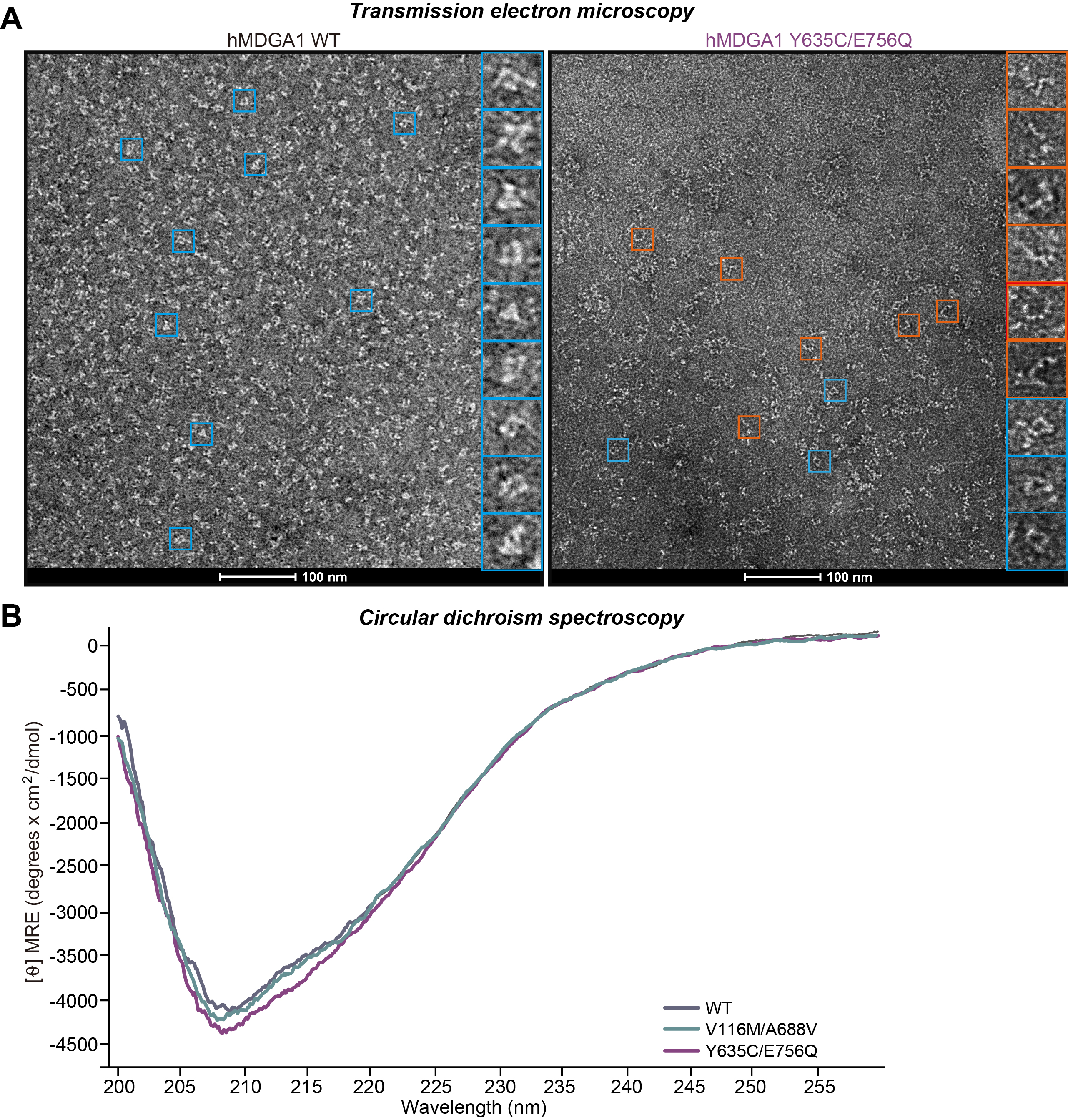

**Supplemental Figure 4. Negative-stain electron microscopy of the full ecto-domain of hMDGA1 WT and hMDGA1 Y635C/E756Q protein.**

(**A**) Representative negative-stained electron microscopy images of the full ectodomain of human MDGA1 WT (**left**) and Y635C/E756Q mutant (**right**). The right insets show close-up views corresponding to the square boxes in the raw micrographs. The closed triangular shape of MDGA1 WT/mutant and the linear shape of MDGA1 mutant are indicated with blue and red boxes, respectively. Scale bar, 100 nm.

(**B**) Far-UV CD spectra of MDGA1 WT and the ASD-associated MDGA1 mutant proteins. The CD spectra are similar, indicating that the secondary structure content of the MDGA1 variants is comparable to that of MDGA1 WT.

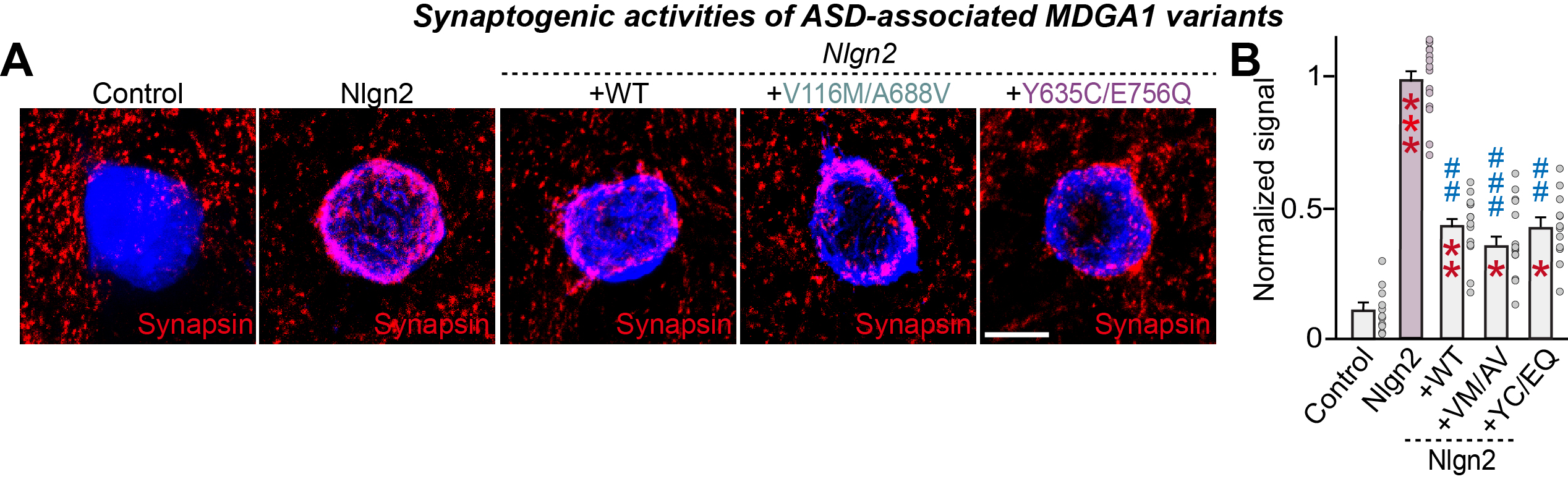
 **Supplemental Figure 5. Analysis of the synapse-suppressing activities of the ASD-associated MDGA1 variants.**

(**A** and **B**) Effects of MDGA1 WT or its variants on the synaptogenic activities of Nlgn2. HEK293T cells expressing the indicated proteins were co-cultured with hippocampal neurons without or with coexpression of the indicated MDGA1 WT or its splice variants. Representative images (**a**) of co-cultures immunostained with antibodies against EGFP or HA (blue) and synapsin I (red). Scale bar, 10 μm (applies to all images). Quantitation (**b**) of heterologous synapse-formation assays, determined by calculating the ratio of synapsin to EGFP/HA fluorescence signals. Data are means ± SEMs (**p* < 0.05, ***p* < 0.01, ****p* < 0.001; nonparametric Kruskal-Wallis test with Dunn’s *post hoc* test; n = 11–14 cells/group).

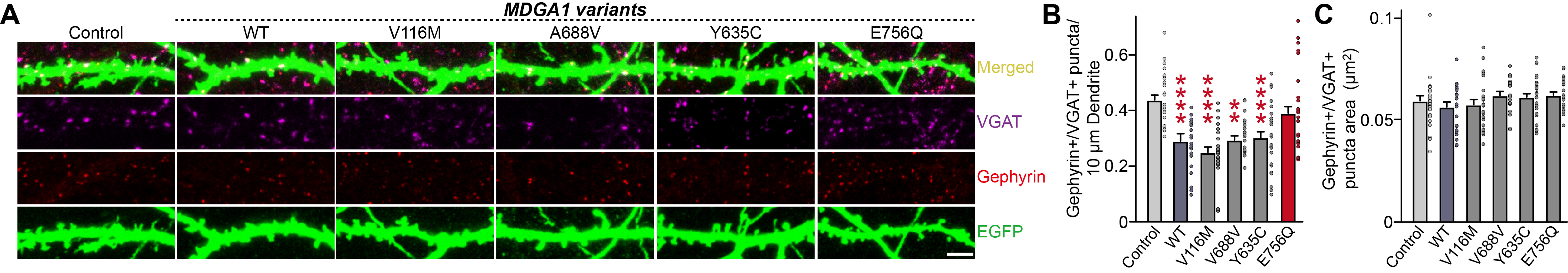

**Supplemental Figure 6. Effects of overexpression of the ASD-associated MDGA1 variants on GABAergic synapses in cultured hippocampal neurons.**

(**A**) Representative images of hippocampal neurons transfected at DIV7 with MDGA1 WT and the indicated MDGA1 variants (V116M, Y635C, A688V and E756Q). Neurons were immunostained at DIV14 for VGAT (magenta) and gephyrin (red). EGFP (green) marks the transfected neurons. Scale bar, 10 µm.

(**B**) Quantification of gephyrin^+^/VGAT^+^ puncta density in hippocampal neurons expressing control, MDGA1 WT or the indicated MDGA1 variant. Data are presented as the number of gephyrin^+^/VGAT^+^ puncta per 10 µm of dendrite. Data are presented as means ± SEMs (n = 19–36 neurons/group; ***p* < 0.01, *****p* < 0.0001; nonparametric Kruskal-Wallis test with Dunn’s *post hoc* test).

(**C**) Quantification of gephyrin^+^/VGAT^+^ puncta area in hippocampal neurons expressing control, MDGA1 WT or the indicated MDGA1 variant. Data are presented as means ± SEMs (n = 19–36 neurons/group).

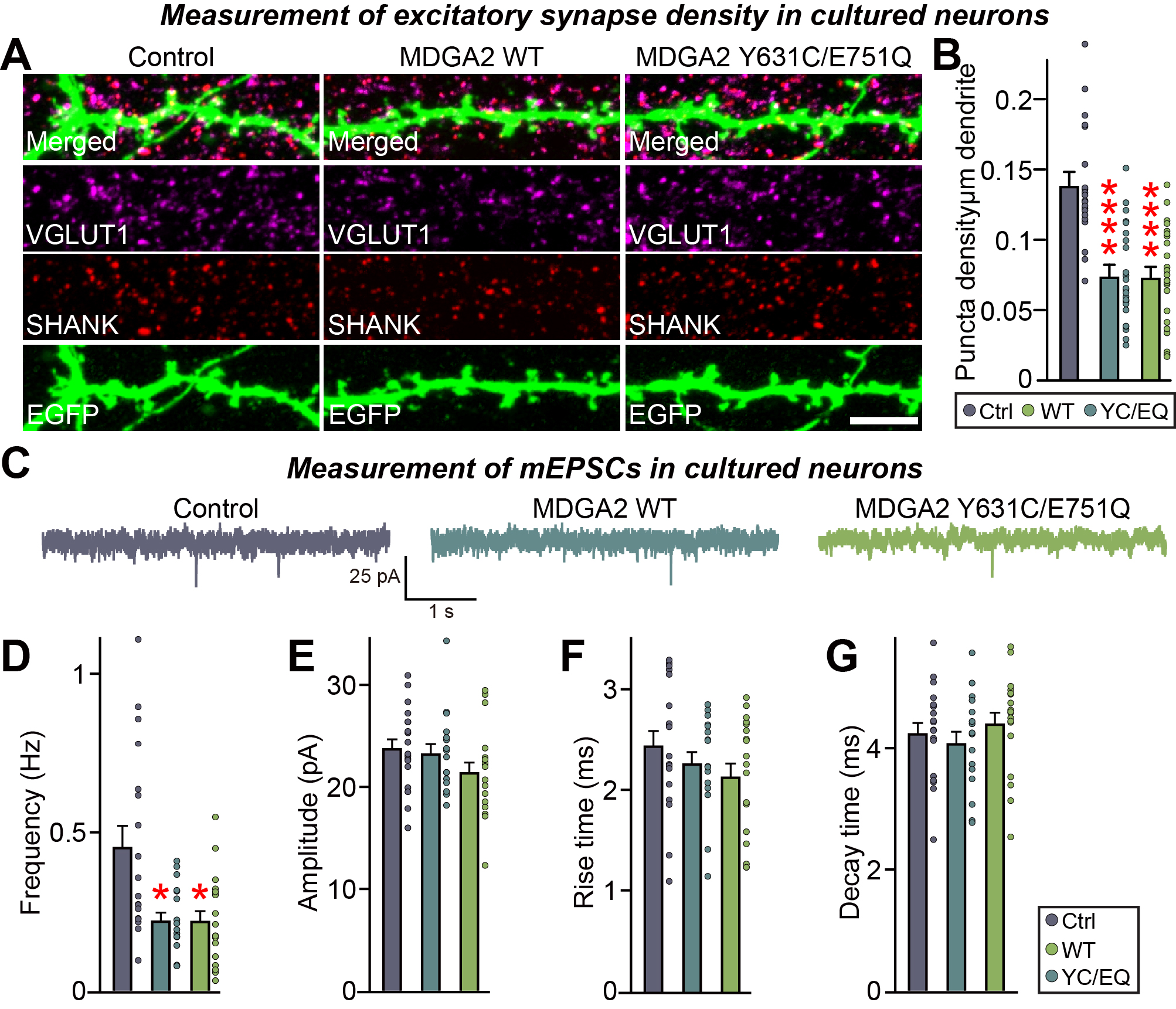

**Supplemental Figure 7. Effects of overexpression of MDGA2 variants (equivalent MDGA1 residues associated with ASDs were introduced) on glutamatergic synapses in cultured hippocampal neurons.**

(**A**) Representative images of hippocampal neurons transfected with MDGA2 WT and MDGA2 Y631C/E751Q mutations. Neurons were immunostained for VGLUT1 (magenta) and Shank (red). EGFP (green) marks the transfected neurons. Merged images show the colocalization of VGLUT1 and Shank. Scale bar, 10 µm.

(**B**) Quantification of VGLUT1^+^/Shank^+^ puncta density in control, MDGA2 WT, and MDGA2 Y631C/E751Q-transfected hippocampal neurons. Data are presented as the number of VGLUT1^+^/Shank^+^ puncta per 10 µm of dendrite. Data are presented as means ± SEMs (n = 21–25 neurons/group; **** *p* < 0.0001; nonparametric Kruskal-Wallis test with Dunn’s *post hoc* test).

(**C–G**) Representative mEPSCs traces (**C**) and quantification of frequency (**D**), amplitude (**E**), rise time (**F**) and decay time (**G**) of mEPSCs from cultured neurons transfected with MDGA2 WT or its variants. Data are presented as means ± SEMs (n = 16–19 neurons/group; **p* < 0.05; ANOVA with a nonparametric Kruskal-Wallis test).

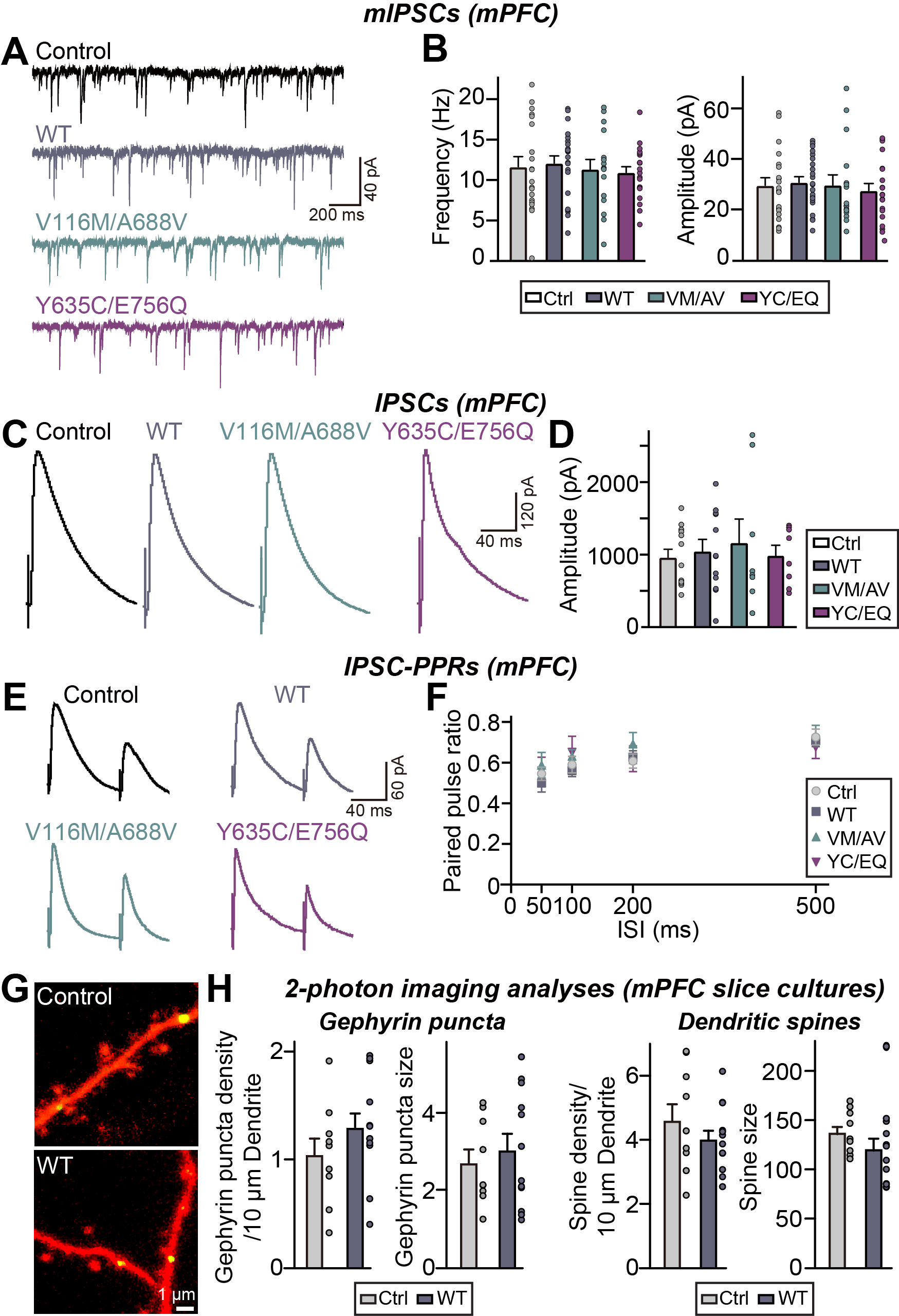

**Supplemental Figure 8. Effects of overexpression of the ASD-associated MDGA1 variants on GABAergic synapses in adult mPFC layer II/III pyramidal neurons.**

(**A** and **B**) Whole-cell recordings of mIPSCs from adult mPFC layer II/III pyramidal neurons expressing control, MDGA1 WT, MDGA1 V116M/A668V or MDGA1 Y635C/E756Q. Representative traces (**A**) and averages of mIPSC frequencies and amplitudes (**B**; control, n = 20/5; WT, n = 22/5; V116M/A668V, n = 16/4; Y635C/E756Q, n = 20/5, where ‘n’ denotes the number of cells/mice). Data are presented as means ± SEMs.

(**C** and **D**) Recordings of eIPSCs from adult mPFC layer II/III pyramidal neurons expressing control, MDGA1 WT, MDGA1 V116M/A688V or MDGA1 Y635C/E756Q. Representative traces (**C**) and average of eIPSC amplitudes (**D**; control, n = 13/4; WT, n = 12/4; V116M/A668V, n = 8/3; Y635C/E756Q, n = 8/3). Data are presented as means ± SEMs.

(**E** and **F**) Recordings of eIPSC-PPRs from adult hippocampal CA1 pyramidal neurons expressing control, MDGA1 WT, MDGA1 V116M/A688V or MDGA1 Y635C/E756Q. Representative traces (**E**) and average of eIPSC-PPRs (**F**; control, n = 13/4; WT, n = 12/4; V116M/A668V, n = 8/3; Y635C/E756Q, n = 8/3). Data are presented as means ± SEMs (**p* < 0.05, ***p* < 0.01, ****p* < 0.001; Kruskal–Wallis test followed by Dunn’s multiple comparison test).

(**G**) Two-photon images of dendritic segments from mPFC layer II/III pyramidal neurons co-transfected with tdTomato, gephyrin intrabody-GFP, and control or MDGA1 WT plasmid.

(**H**) Quantitative analysis of gephyrin^+^ puncta density and size (**left**) and dendritic spine density and size (**right**).

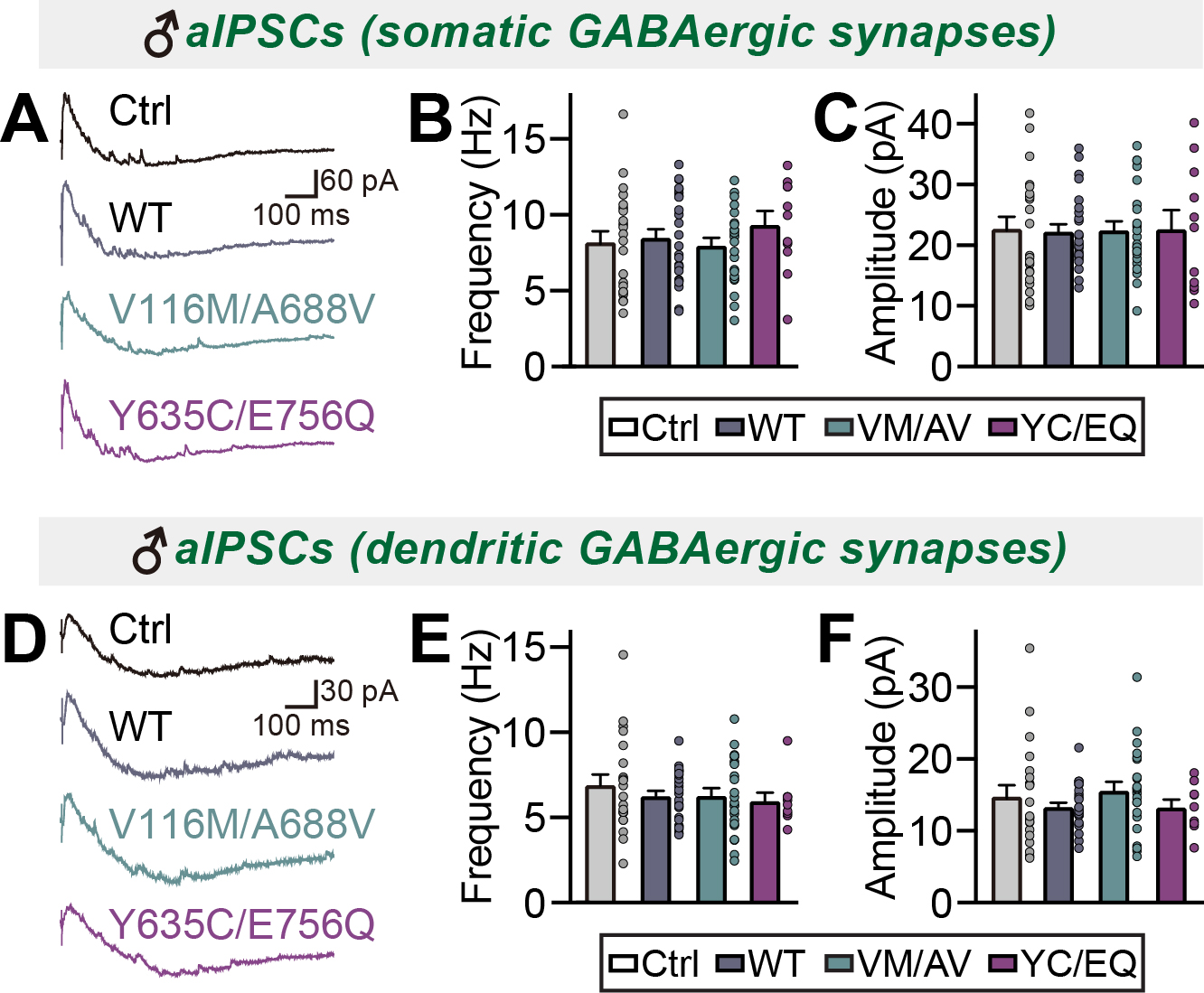

**Supplemental Figure 9. Effects of overexpression of the ASD-associated MDGA1 variants on asynchronous evoked GABAergic transmission in adult hippocampal CA1 pyramidal neurons.**

(**A–C**) Representative traces (**A**) and averages of somatic aIPSC frequencies (**B**) and amplitudes (**C**) from CA1 pyramidal neurons (Control, n = 22/5; WT, n = 25/5; V116M/A668V, n = 23/5; Y635C/E756Q, n = 11/4; ‘n’ denotes number of cells/mice).

(**D–F**) Representative traces (**D**) and averages of dendritic aIPSC frequencies (**E**) and amplitudes (**F**) from CA1 pyramidal neurons (Control, n = 21/5; WT, n = 25/5; V116M/A668V, n = 23/5; Y635C/E756Q, n = 9/4). Paired electric stimulation at 10 Hz was used for measuring aIPSCs. Data are presented as means ± SEMs (Kruskal–Wallis test followed by Dunn’s multiple comparison test).

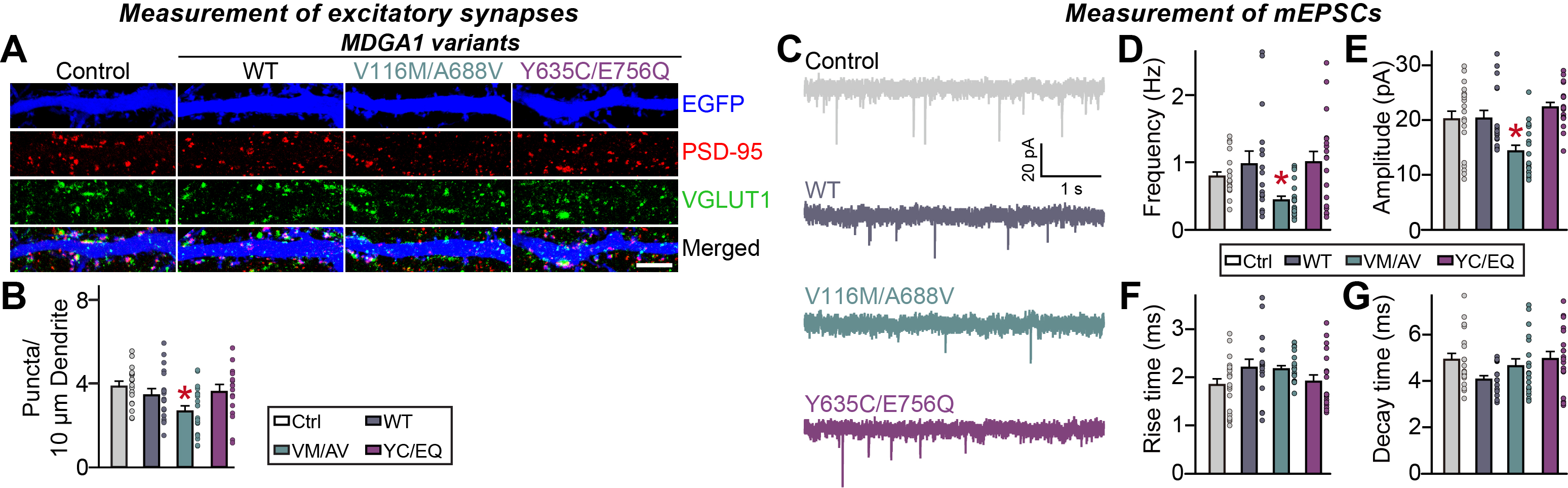
**Supplemental Figure 10. Effects of overexpression of the ASD-associated MDGA1 variants on glutamatergic synapses in cultured hippocampal neurons.**

(**A** and **B**) Representative images (**A**) and summary graphs (**B**) showing the density of glutamatergic synaptic puncta of cultured hippocampal neurons transfected at DIV7 with the indicated full-length MDGA1 expression construct and immunostained at DIV14 with antibodies to VGLUT1 and PSD-95 (excitatory synaptic markers) and EGFP. Data are presented as means ± SEMs (**p* < 0.05; ANOVA with non-parametric Kruskal-Wallis test; n = 17–21 images/group). Scale bar, 10 μm (applies to all images).

(**C–G**) Representative mEPSCs traces (**C**) and quantification of frequency (**D**), amplitude (**E**), rise time (**F**) and decay time (**G**) of mEPSCs from cultured neurons transfected with MDGA1 or its variants. Data are presented as means ± SEMs (n = 16–21 neurons/group; **p* < 0.05; ANOVA with nonparametric Kruskal-Wallis test).

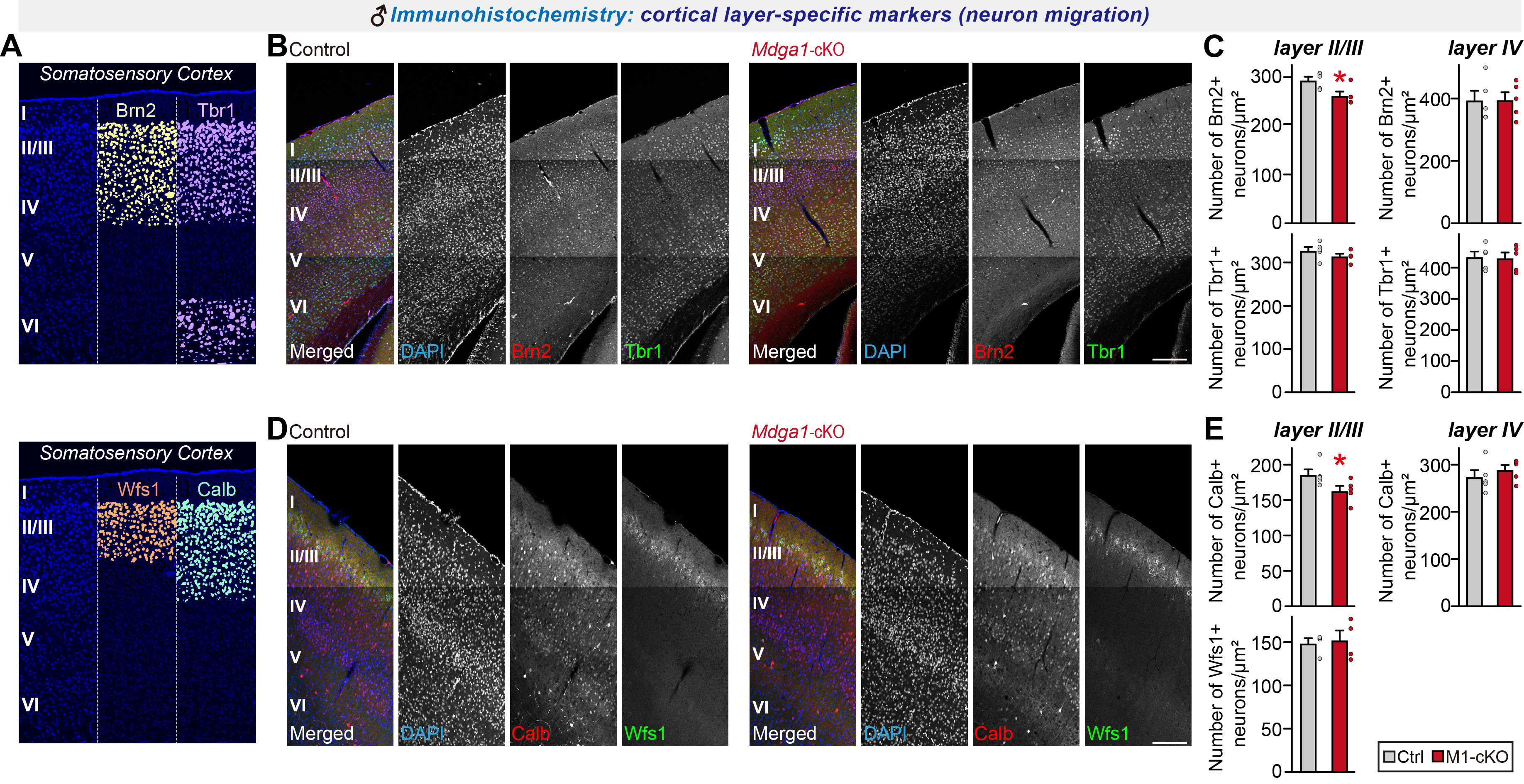

**Supplemental Figure 11. Analysis of cortical neuron migration for adult male *Mdga1*-cKO mice.**

(**A**) Schematic of cortical layer-specific markers used in the current study.

(**B**) Representative images of Brn2- and Tbr1-labeled neurons in the somatosensory cortex of adult male control and *Mdga1*-cKO mice. Neurons were immunostained with antibodies against Brn2 (red), Tbr1 (green) and DAPI (blue). Brn2 and Tbr1 are cortical layer-specific markers (Brn2 for layers II–IV; Tbr1 for layers II/III and primarily VI). Scale bar, 200 μm (applies to all images).

(**C**) Quantification of Brn2^+^ and Tbr1^+^ neuron density in cortical layers (layers II/III and IV) of adult male control and *Mdga1*-cKO mice. Data are presented as means ± SEMs (n = 5 mice/group; **p* < 0.05; Mann–Whitney *U* test).

(**D**) Representative images of calbindin- and Wfs1-labeled neurons in the somatosensory cortex of adult male control and *Mdga1*-cKO mice. Neurons were immunostained with antibodies against calbindin (Calb; red), Wfs1 (green) and DAPI (blue). Calb and Wfs1 are cortical layer-specific markers (Calb for layers II–IV; Wfs1 for layers II/III). Scale bar, 200 μm (applies to all images).

(**E**) Quantification of calbindin^+^ and Wfs1^+^ neuron density in cortical layers II/III and IV of adult male control and *Mdga1*-cKO mice. Data are presented as means ± SEMs (n = 5 mice/group; **p* < 0.05; Mann–Whitney *U* test).

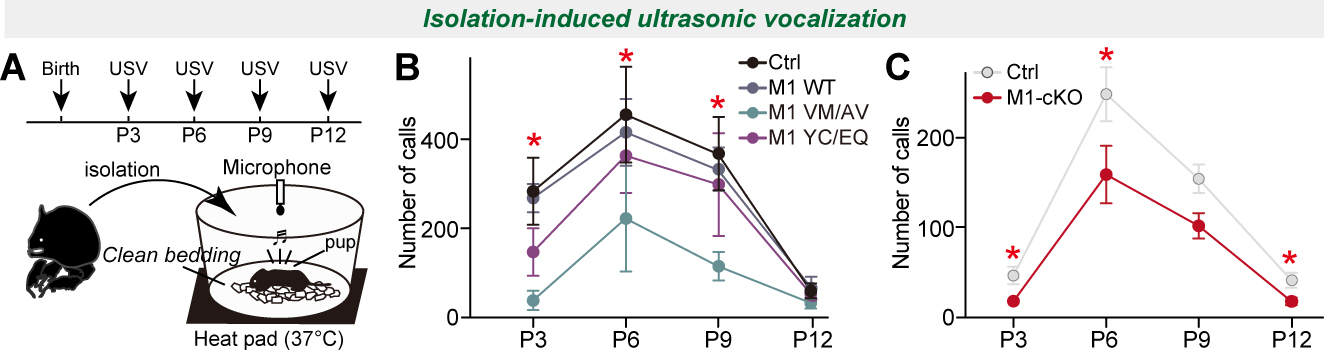
**Supplemental Figure 12. Analysis of isolation-induced ultrasonic vocalizations in mice expressing ASD-associated MDGA1 variants or *Mdga1*-cKO mice.**

(**A**) Experimental setup for recording ultrasonic vocalizations (USVs) from isolated pups. Pups were separated from their mother and placed on clean bedding with a heat pad (37°C). A microphone was used to record the USVs. USV recordings from WT pups transfected in utero with the indicated MDGA1 variants were measured at postnatal days P3, P6, P9 and P12.

(**B**) Number of USV calls recorded from pups expressing the control or the indicated MDGA1 variant at P3, P6, P9 and P12. Data are presented as means ± SEM (n = 7–9 pups/group; asterisks (*) denote significant differences between control and VM/AV group; **p* < 0.05, Mann–Whitney *U* test).

(**C**) Number of USV calls recorded from control and *Mdga1*-cKO pups at P3, P6, P9 and P12. Data are presented as means ± SEMs (n = 19–24 pups/group; **p* < 0.05, Mann–Whitney *U* test).

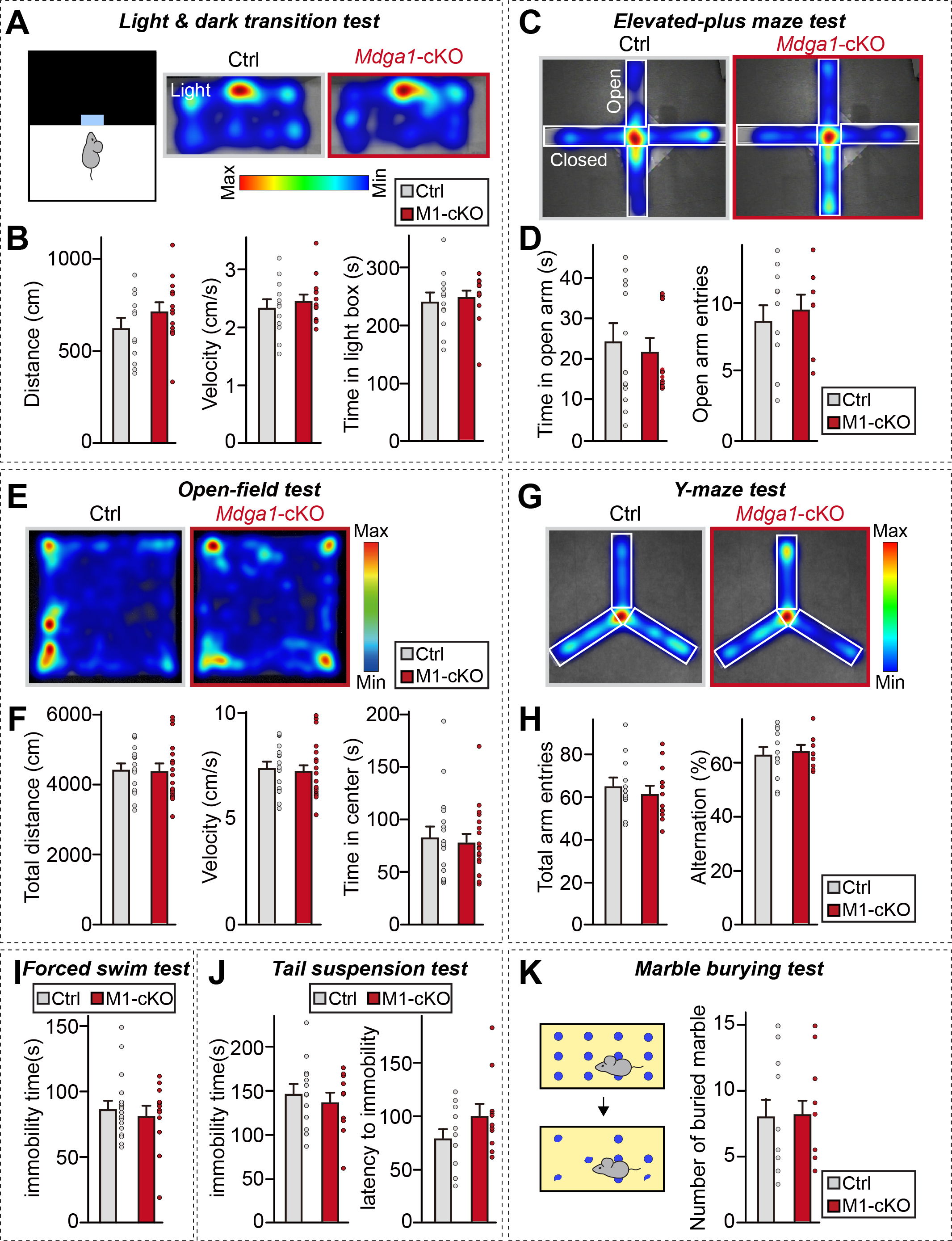
**Supplemental Figure 13. Analysis of behaviors of adult male *Mdga1*-cKO mice.**

(**A** and **B**) Light and dark transition test results showing time spent in the light box, total distance traveled, and velocity for control and *Mdga1*-cKO mice. Data are presented as means ± SEMs (n = 12–14 mice/group).

(**C** and **D**) Elevated-plus maze test results showing time spent in the open arms and number of open arm entries for control and *Mdga1*-cKO mice. Data are presented as means ± SEMs (n = 9–12 mice/group).

(**E** and **F**) Open-field test results showing total distance traveled, time spent in the center, and velocity for control and *Mdga1*-cKO mice. Data are presented as means ± SEMs (n = 17–18 mice/group).

(**G** and **H**) Y-maze test results showing total arm entries and alternation percentage for control and *Mdga1*-cKO mice. Data are presented as means ± SEMs (n = 12 mice/group).

(**I**) Forced swim test results showing total immobility time for control and *Mdga1*-cKO mice. Data are presented as means ± SEMs (n = 11–18 mice/group).

(**J**) Tail suspension test results showing the latency to immobility and total immobility time for control and *Mdga1*-cKO mice. Data are presented as means ± SEMs (n = 11–13 mice/group).

(**K**) Marble burying test results showing the number of marbles buried by control and *Mdga1*-cKO mice. Data are presented as means ± SEMs (n = 13–15 mice/group).

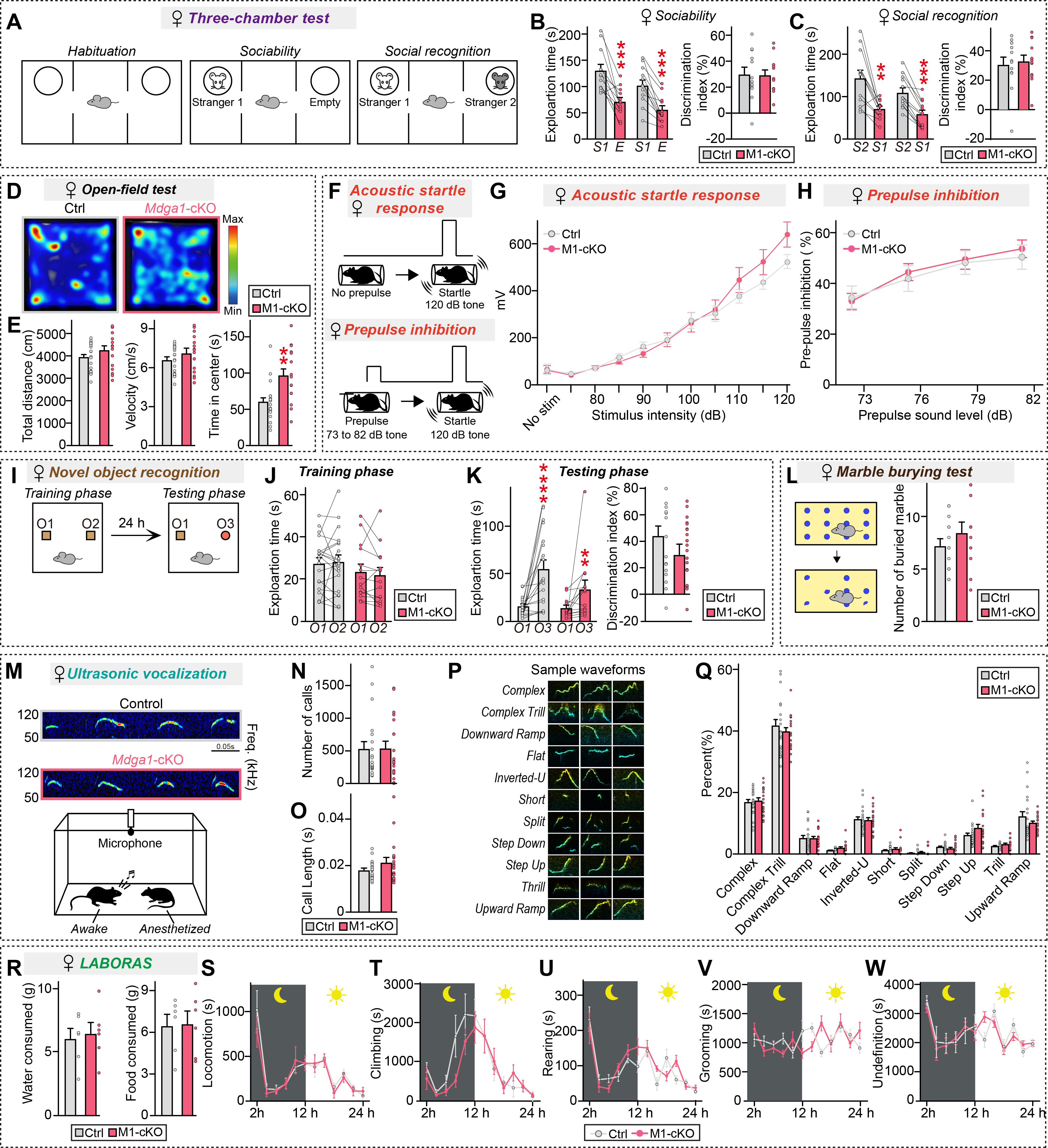

**Supplemental Figure 14. Analysis of behaviors of adult female *Mdga1*-cKO mice.**

(**A–C**) Three-chamber test results showing a schematic of the test (**A**) and the exploration time and discrimination index during the sociability (**B**; stranger 1 vs. empty) and social recognition (**C**; stranger 1 vs. stranger 2) phases for female control and *Mdga1*-cKO mice. Data are presented as means ± SEM (n = 12 mice/group; ***p* < 0.01, ****p* < 0.001; Mann–Whitney *U* test).

(**D** and **E**) Open-field test results showing total distance traveled, velocity, and time spent in the center for female control and *Mdga1*-cKO mice. Data are presented as means ± SEMs (n = 15–20 mice/group; ***p* < 0.01; Mann–Whitney *U* test).

(**F**) Schematic of the prepulse inhibition (PPI) test. Mice were exposed to a prepulse sound (73 to 82 dB) followed by a startle pulse (120 dB), and the inhibition of the startle response was measured.

(**G**) Acoustic startle response in female control and *Mdga1*-cKO mice. Data are presented as means ± SEMs (n = 17–18 mice/group).

(**H**) PPI of the acoustic startle response in female control and *Mdga1*-cKO mice. Data are presented as means ± SEMs (n = 14–17 mice/group).

(**I–K**) Novel object recognition test results showing a schematic of the test (**I**) and the exploration time and discrimination index during the training (**J**) and testing phases (**K**) for female control and *Mdga1*-cKO mice. Data are presented as means ± SEMs (n = 13–18 mice/group; ***p* < 0.01, *****p* < 0.0001; Wilcoxon matched-pairs signed rank test).

(**L**) Marble burying test results showing the number of marbles buried by female control and *Mdga1*-cKO mice. Data are presented as means ± SEMs (n = 11–14 mice/group).

(**M–Q**) Ultrasonic vocalization results showing a representative sonogram (**M**) and the number of calls (**N**) and call length (**O**) for female control and *Mdga1*-cKO mice. Representative waveforms (**P**) and distribution of different call types (**Q**) are shown. Data are presented as means ± SEMs (n = 18–20 mice/group; Mann–Whitney *U* test).

(**R–W**) LABORAS test results showing various homecage behaviors, water and food consumption (**R**), locomotion (**S**), climbing (**T**), rearing (**U**), grooming (**V**), and undefined activities (**W**) for female control and *Mdga1*-cKO mice measured over 24 hours. Data are presented as means ± SEMs (n = 6 mice/group).

**Supplemental Table 1. Description of patients with the ASD-associated *MDGA1* variants described in the current study and the functional effects of these variants predicted using four different bioinformatics tools**

| **Variant** | **First Family** | | | | **Second Family** | | | |
| --- | --- | --- | --- | --- | --- | --- | --- | --- |
| Position | g.37612408C>G | | g.37615091T>C | | g.37626057C>T | | g.37614135G>A | |
| DNA change | c.2266G>C | | c.1904A>G | | c.346G>A | | c.2063C>T | |
| Amino acid change | p.Glu756Gln | | p.Tyr635Cys | | p.Val116Met | | p.Ala688Val | |
| Exon | 13 | | 15 | | 3 | | 11 | |
| ACMG Classification | Uncertain Significance | | Uncertain Significance | | Uncertain Significance | | Likely Benign | |
| Gnomad Frequency | ≈0.07% | | ≈0.001% | | <0.001% | | ≈0.07% | |
| Healthy Homozygotes | No | | No | | No | | No | |
| CADD score | 32 | | 27.2 | | 23.6 | | 21.3 | |
| SIFT | Damaging | Damaging | |  | | Damaging | | Tolerated |
| FATHMM-MKL | Damaging | Damaging | |  | | Damaging | | Damaging |
| Eigen | Pathogenic | Pathogenic | |  | | Pathogenic | | Benign |
| LRT | Deleterious | Deleterious | |  | | Deleterious | | Deleterious |
| Mutation Taster | Disease causing | Disease causing | |  | | Disease causing | | Neutral |
| MVP | Benign | Benign | |  | | Benign | | Benign |

Abbreviations: N.D., not determined

**Supplemental Table 2. Gene set enrichment test for cluster association with neurological disorders**

**Supplemental Table 3. Stability analysis of ASD-associated *MDGA1* mutations**

The free energy of folding (stability) was calculated using FoldX5^92^

| **Model** | **Energy (kcal.mol-1)** |
| --- | --- |
| MDGA1 (Q19-P742) WT | 0.04 |
| MDGA1 (Q19-P742) V116M | 0.04 |
| MDGA1 (Q19-P742) A688V | 1.89 |
| MDGA1 (Q19-P742) Y635C | 7.02 |

Abbreviations: WT, wild type
